# Cohort Profile Update: Expanding the Cardiovascular Risk in Young Finns Study into a multigenerational cohort

**DOI:** 10.1101/2024.11.24.24317769

**Authors:** Katja Pahkala, Suvi Rovio, Kari Auranen, Matthieu Bourgery, Marko Elovainio, Mikael Fogelholm, Johanna Haapala, Mirja Hirvensalo, Nina Hutri, Eero Jokinen, Antti Jula, Markus Juonala, Jari Kaikkonen, Noora Kartiosuo, Hannu Kiviranta, Juhani S. Koskinen, Noora Kotaja, Mika Kähönen, Tomi P. Laitinen, Terho Lehtimäki, Irina Lisinen, Britt-Marie Loo, Leo-Pekka Lyytikäinen, Costan G. Magnussen, Pashupati P. Mishra, Juha Mykkänen, Juho-Antti Mäkelä, Satu Männistö, Jaakko Nevalainen, Laura Pulkki-Råback, Emma Raitoharju, Panu Rantakokko, Tapani Rönnemaa, Sini Stenbacka, Leena Taittonen, Tuija H. Tammelin, Jorma Toppari, Päivi Tossavainen, Jorma Viikari, Olli Raitakari

## Abstract

Cardiovascular Risk in Young Finns Study (YFS) is a prospective cohort of 3596 males and females (baseline age 3-18 years) that was established in 1980 to study cardiovascular risk factors in children and adolescents. The YFS has been instrumental in demonstrating the links between childhood risk factors and adult cardiovascular outcomes with implications on paediatric cardiovascular preventive practise.

In the latest follow-up in 2018-2020, the study was expanded into a three-generation cohort, including the original cohort members, as well as their parents and offspring. Altogether 7341 individuals aged 3-92 years participated; 2127 original cohort members (now aged 40-58 years), 2452 parents (aged 58-92 years) and 2762 offspring (aged 3-37 years) of the original cohort members.

The main aim was to establish a multigenerational population data base, sample biobank and links to national health registries that would offer a unique platform to study familial transmission of health-related traits. The field studies and data collections were conducted as part of the ERC funded MULTIEPIGEN project designed testing a specific hypothesis that epigenetic markers in the semen play a role in the transmission of intergenerational information in the paternal lineage. It is a first *a priori* designed epidemiologic study assessing the role of paternal life exposures, including chemical and psychosocial stress, in the development of cardio-metabolic, cognitive and psychosocial outcomes in their offspring.

## The original cohort

The long-standing national multi-centre Cardiovascular Risk in Young Finns Study (YFS) was originally designed to provide information on cardiovascular risk factors and their determinants in children and adolescents of various ages in different parts of Finland.(1,2) The first examination in 1980 recruited 3596 participants aged 3-18 years from five cities and their surrounding rural communities. The cohort has been followed up 8 times with large-scale field studies in 1983, 1986, 1989, 1992, 2001, 2007, 2011, and 2018-2020 The participation rates have varied between 61 and 85%. In the latest follow-up study, the recruitment was extended to cover three consecutive generations: the original participants as well as their parents and offspring.

At present, the YFS is the largest study in Europe with a follow-up of cardiometabolic and other health-related phenotypes from childhood to adulthood. With over 1200 publications, the study has contributed to understanding the development of cardiometabolic risk, including diet, lifestyle, metabolic risk factors, psychological traits and psychosocial factors, inflammation, hormones and genetic markers. Although the primary focus has been on cardiometabolic health research, the data are increasingly being utilised to examine many other aspects of health and well-being.

The YFS is part of the International Childhood Cardiovascular Cohort (i3C) Consortium, which has collected prospective data from 38,589 children in all major cardiovascular cohorts that follow participants from childhood to adulthood to examine the relationships between childhood health and adult cardiovascular outcomes.(3–5) In addition, by leveraging its extensively phenotyped data, the YFS investigators continually contribute to international collaborative studies.(6,7)

## What is the reason for the new focus (or new data collection)

In addition to the continued collection of follow-up data, the latest YFS field study was designed to establish a multigenerational cohort that serves as a platform to examine intergenerational transmission of non-communicable diseases in humans. Data from animal models indicate that exposure to various stressors can lead to phenotypic changes not only in the predisposed individuals but also in future generations, such that individuals can acquire phenotypes caused by exposures of their ancestors. Such effects may operate with epigenetic mechanisms that do not involve new DNA mutations. In humans, it is not possible to perform multigenerational controlled trials to examine epigenetic mechanism, so it has been suggested that epidemiologic studies should be reframed to include exposures from previous generations to examine the mechanisms that transmit this information to offspring.(8) Therefore, the data collections in the most recent follow-up field study were accomplished as part of a ERC funded project entitled MULTIEPIGEN (Ancestral environmental exposures and offspring health – a multigenerational epidemiologic cohort study across 3 generations), which is the first *a priori* designed epidemiologic study assessing the role of ancestral exposures influencing offspring health (multiepigen.utu.fi/).

## What will be the new areas of research?

Our general aim was to establish a multigenerational population database, sample biobank and to link these to national health registries, providing a unique platform to examine the role of epigenetic inheritance and others forms of familial transmission in human health. The novel research areas comprise the study of epigenetic transmission in humans, which allows the investigation of links between parental exposures and offspring phenotypes. The pre-defined ancestral exposures include tobacco smoke, environmental toxicants, and accumulation of psychosocial adversities, which based on prior experimental evidence have a very high plausibility to cause intergenerational effects on obesity-related phenotypes, cognitive function and psychological wellbeing. The establishment of a multigenerational epidemiologic cohort allows us to explore whether parental exposure to these extrinsic and intrinsic stressors is related to the offspring’s characteristics.

The sample and data collection during the field study were comprehensive and included e.g. whole-blood, serum and semen to allow measurement of genetic architecture, transcriptomics, and epigenetic markers, including DNA methylation and non-coding RNA molecules. Such broad-scope data collection was justified because it is unknown how the epigenetic mechanisms operate in humans. It is thus crucial to keep an open mind to the potential mechanisms that transmit this information to offspring. Specifically, this new research area will test the hypothesis that non-coding RNA molecules and DNA methylation in the sperm play a role in the transmission of intergenerational information in the paternal lineage in humans, as has been previously shown in rodents.(9) The detailed statistical analysis plan to test the original hypotheses listed in the research plan of the MULTIEPIGEN project (https://cordis.europa.eu/project/id/742927) is provided in the Supplement.

## Who is in the cohort?

The data collection included the original YFS participants (generation 1, G1) as well as their parents (generation 0, G0) and offspring (generation 2, G2; age ≥3 years). This first field study of the multigenerational cohort was executed from March 2018 to February 2020. Of the original 3596 G1 participants, 137 had died and 150 had withdrawn from the study. Consequently, we contacted the remaining G1 participants (n=3309) to inform them about the beginning of the new multigenerational data collection. At this time, they had the possibility to restrain us from inviting their parents and/or offspring. After excluding those who had deceased, decided to withdraw from the study or for whom we had no contact information, we invited 3217 G1 participants. Additionally, we invited 3940 G0 participants and 5696 G2 participants, including parents and adult offspring of those G1 participants who had deceased. In total, the number of invited individuals was 12853. Altogether 7341 of them provided any data (57.1%). The number of participants who attended the study visit was 6753 (52.5%) while 588 participants provided only questionnaire data. The generation-specific participation rates for participants with any data were 66.1% for G1 (N=2127; females 55.0%; age 40-58 years), 62.2% for G0 (N=2452; females 61.1%; age 58-92 years), and 48.5% for G2 (N=2762; females 54.9%; age 3-37 years). Figure 1 illustrates the recruitment process. Of those participants who attended the study visit, 6214 (92.0%) attended after no/minimal reminding, 283 (4.2%) after the first re-invitation letter, and 256 (3.8%) after additional invitations.

**Figure 1.**
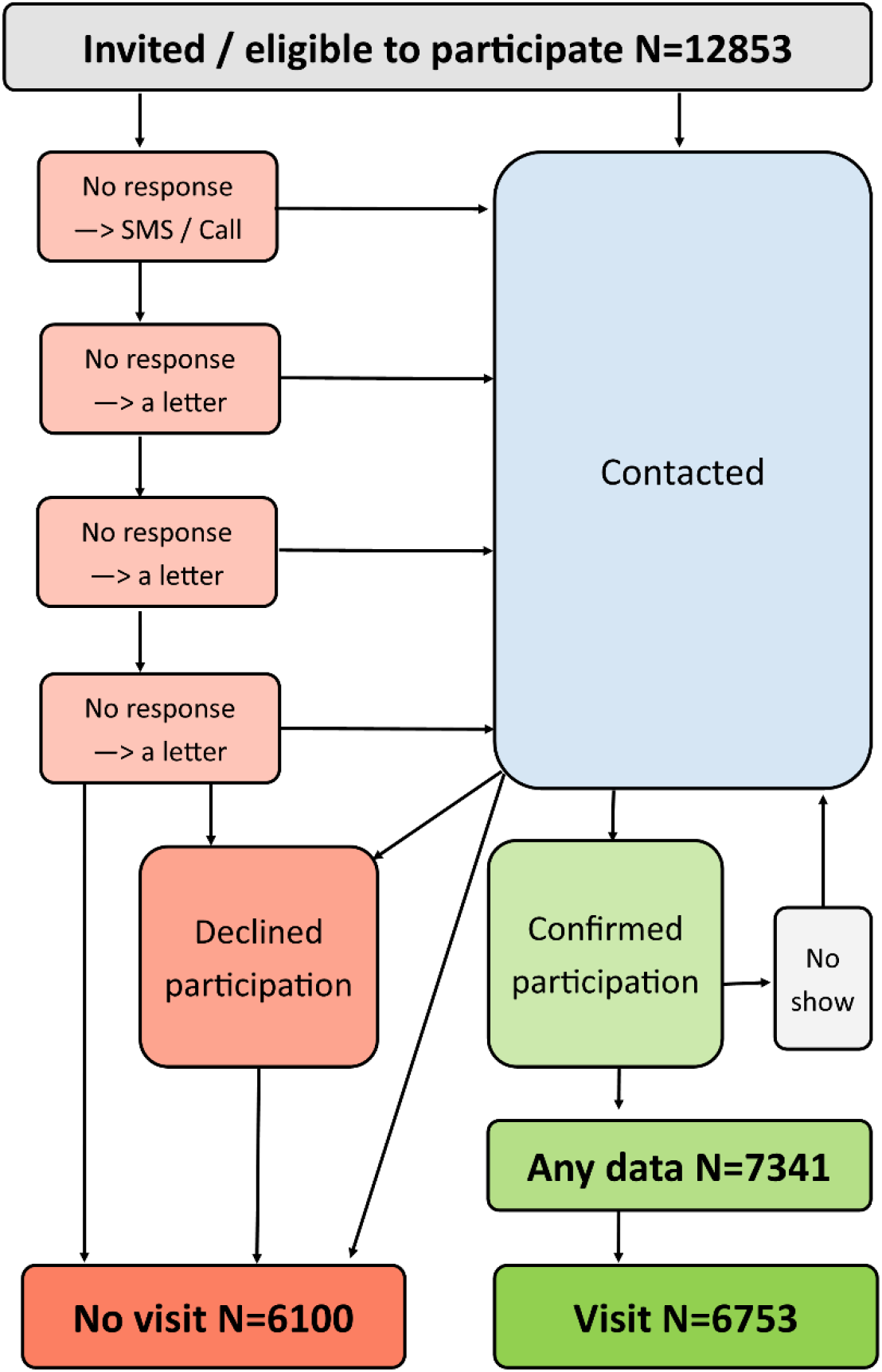
Recruitment process in the field study.

Stemming from the multigenerational aspect, the participants form 2385 triads (1956 attended study visit) including data from all three generations (G0-G1-G2; Figure 2). Dyads – pairs of a parent and an offspring – are provided by 1879 G0-G1 participant dyads (1656 attended study visit; one G1 participant may be included in one or two dyads based on the number G0 participants) and 2499 G1-G2 participant dyads (2295 attended study visit; one G1 participant may be included in several dyads based on the number of related G2 participants). Additionally, the cohort included 135 G0-G2 dyads (99 attended study visit).

**Figure 2.**
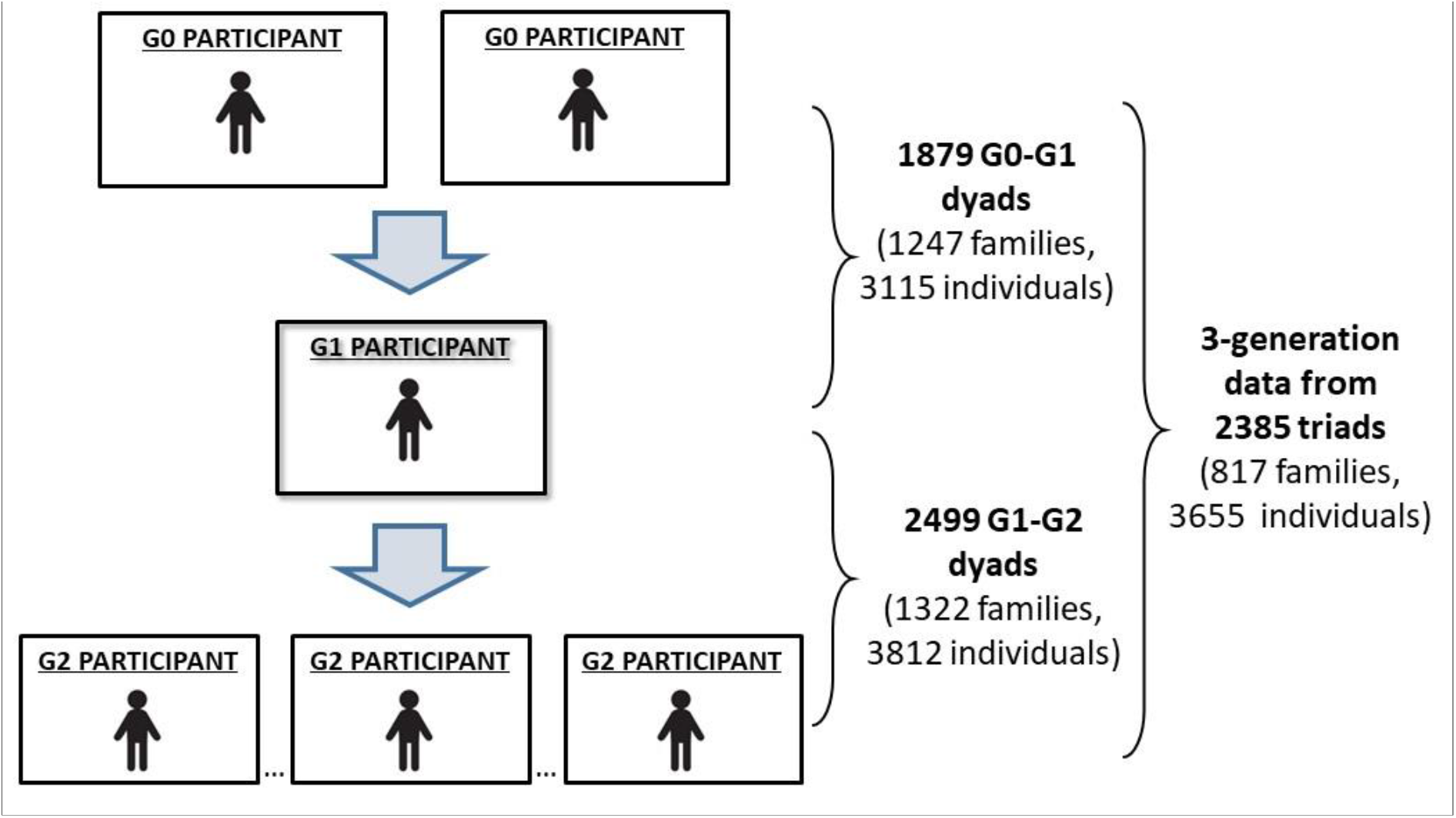
Three-generational study design of the MULTIEPIGEN project.

## What has been measured?

The assessment timeline of some key measures collected in YFS is presented in Table 1. Related to the specific research aims of the MULTIEPIGEN project, we assessed detailed data on the ancestral exposures, i.e. tobacco smoke, environmental toxicants, and the accumulation of psychosocial adversities at the time of conception and pregnancy. Regarding tobacco smoke, the exposure was additionally assessed in early childhood (age 0-6 years) and childhood/adolescence (age 7-18 years). To obtain data on exposure to tobacco smoke, we used detailed prospective and retrospective questionnaires and the measurement of serum cotinine. The assessed environmental toxicants include serum concentrations of polychlorinated biphenyls, organochlorine pesticides and their metabolites, polybrominated diphenyl ethers and perfluorinated alkyl substances. Detailed questionnaires were used to collect data on psychosocial exposures, e.g. measures of socioeconomic factors, emotional factors, parental health behaviours, stressful life events, self-regulation of the child, and social adjustment of the child.

**Table 1.** Examples of collected data in the YFS.

| Generation |  |  |  |  |  |  | MULTIEPIGEN |  |  |
| --- | --- | --- | --- | --- | --- | --- | --- | --- | --- |
|  |  |  |  |  |  |  | G1 | G0 | G2 |
|  | Longitudinal data (original YFS cohort) |  |  |  |  |  |  |  |  |
| Data from YFS follow-ups | 1980 | 1983 | 1986 | 2001 | 2007 | 2011 | 2018 | 2018 | 2018 |
| Age of participants (years) | 3-18 | 6-21 | 9-24 | 24-39 | 30-45 | 34-49 | 41-56 | 59-92 | 3-37 |
| Number of participants | 3596 | 2991 | 2579 | 2284 | 2204 | 2063 | 2127 | 2452 | 2762 |
| Anthropometric measurements |  |  |  |  |  |  |  |  |  |
| Height and weight | x | x | x | x | x | x | x | x | x |
| Waist and hip circumferences<sup>a</sup> |  |  |  | x | x | x | x | x | x |
| Skinfold thicknesses | x | x | x |  |  |  |  |  |  |
| Blood pressure | x | x | x | x | x | x | x | x | x |
| Biochemical markers |  |  |  |  |  |  |  |  |  |
| Lipids and apolipoproteins A1 and B | x | x | x | x | x | x | x | x | x |
| Fatty acids | x | x | x | x |  |  |  |  |  |
| Insulin | x | x | x | x | x | x | x | x | x |
| Glucose |  |  | x | x | x | x | x | x | x |
| C-reactive protein | x |  |  | x | x | x | x |  |  |
| Inflammatory cytokines |  |  |  |  | x |  |  |  |  |
| Cotinine | x | x | x | x | x | x | x |  |  |
| Persistent organic pollutants | x |  |  | x |  |  |  |  |  |
| Metabolomics |  |  |  | x | x | x | x | x | x |
| Lipidomics |  |  |  | x | x | x |  |  |  |
| Urine metabolomics |  |  |  |  |  |  | x | x | x |
| Questionnaire data |  |  |  |  |  |  |  |  |  |
| Physical activity<sup>b</sup> | x | x | x | x | x | x | x | x | x |
| Dietary assessment | x | x | x | x | x | x | x | x | x |
| Smoking (passive/active) | x | x | x | x | x | x | x | x | x |
| Alcohol consumption | x | x | x | x | x | x | x | x | x |
| Psychological well-being | x | x | x | x | x | x | x | x | x |
| Socioeconomic status | x | x | x | x | x | x | x | x | x |
| Ultrasound imaging data |  |  |  |  |  |  |  |  |  |
| Carotid artery IMT and plaque |  |  |  | x | x |  | x | x | x |
| Carotid artery elasticity |  |  |  | x | x |  | x | x | x |
| Brachial artery FMD |  |  |  | x | x |  | x |  |  |
| Cardiac structure and function |  |  |  |  |  | x |  |  |  |
| Coronary calcification |  |  |  |  | x |  |  |  |  |
| Fatty liver |  |  |  |  |  | x | x | x | x |
| **Cognitive function** |  |  |  |  |  |  |  |  |  |
|  |  |  |  |  |  | x | x | x | x |
| **Genetic data** |  |  |  |  |  |  |  |  |  |
| Whole-genome data/GWAS | x |  |  |  |  |  |  | x | x |
| Mitochondrial genome | x |  |  |  |  |  |  |  |  |
| Blood cell transcriptomics |  |  |  |  |  | x |  |  |  |
| Faecal metagenomics |  |  |  |  |  |  | x | x | x |
| **Epigenetic data** |  |  |  |  |  |  |  |  |  |
| Blood cell DNA methylation |  |  | x |  |  | x | x | x | x |
| Blood cell microRNAs |  |  |  |  |  | x |  |  |  |
| Serum microRNAs |  |  |  | x |  | x | x |  |  |
| Sperm small RNAs |  |  |  |  |  |  | x | x | x |
| Sperm DNA methylation |  |  |  |  |  |  | x | x | x |

<sup>a</sup> Hip circumference was not measured in 2018.
<sup>b</sup> In addition to questionnaires, device-measured physical activity was assessed in 2007, 2011 and 2018.

The pre-specified offspring outcomes comprised obesity-related phenotypes, early vasculopathy indicative of subclinical atherosclerosis, cognitive function and psychological well-being. The obesity-related phenotypes included components of the metabolic syndrome, such as measures of adiposity, insulin/glucose metabolism, serum lipoproteins, blood pressure, and fatty liver (assessed with ultrasound). The measures of vascular changes indicative of early vasculopathy include carotid artery plaques, carotid artery intima-media thickness, and carotid artery elasticity, measured using ultrasound. Related to cognitive function, we measured episodic memory and associative learning, working memory, executive function, reaction and movement time, and sustained attention. Outcome data on psychological well-being were collected with questionnaires and included personality characteristics, social relations, depression and anxiety, psychosocial work environment and temperament characteristics.

Additionally, we have collected hair samples to objectively detect stress-related biomarkers such as cortisol concentrations.

To investigate the hypothesis that non-coding RNA molecules and DNA methylation in the sperm mediate transmission of intergenerational epigenetic information in the paternal lineage, we collected semen samples from adult male participants and purified spermatozoa from the semen samples. Sperm total RNA was extracted for small-RNA-sequencing analysis, and genomic DNA was extracted for DNA methylation analysis by Reduced Representation Bisulfite Sequencing. Furthermore, the data collections included several other measurements. A detailed description of all measurements is provided in the Supplement.

During the field study, we applied a thorough reminding protocol to assess as complete data collections as possible regarding the questionnaires, semen and faecal samples (illustrated in Figure 3). Briefly, phone calls and text messages were utilised to increase the final yield of questionnaires and samples among the participants who had attended the study visit. We primarily contacted participants with only one missing questionnaire or sample. We especially focused on 1) forming as complete three-generational family structure as possible, 2) acquiring at least the basic questionnaire from those who had attended the study visit, and 3) acquiring dietary data (i.e., food frequency questionnaire) from the participants who had given a faecal sample. We also offered new study visits to those who had not completed all of the examinations during their primary study visit. Because the semen samples and basic questionnaires played a key role in regard to the research aims of the MULTIEPIGEN project, we contacted once more all participants who had not returned these data after the field study period had ended.

**Figure 3.**
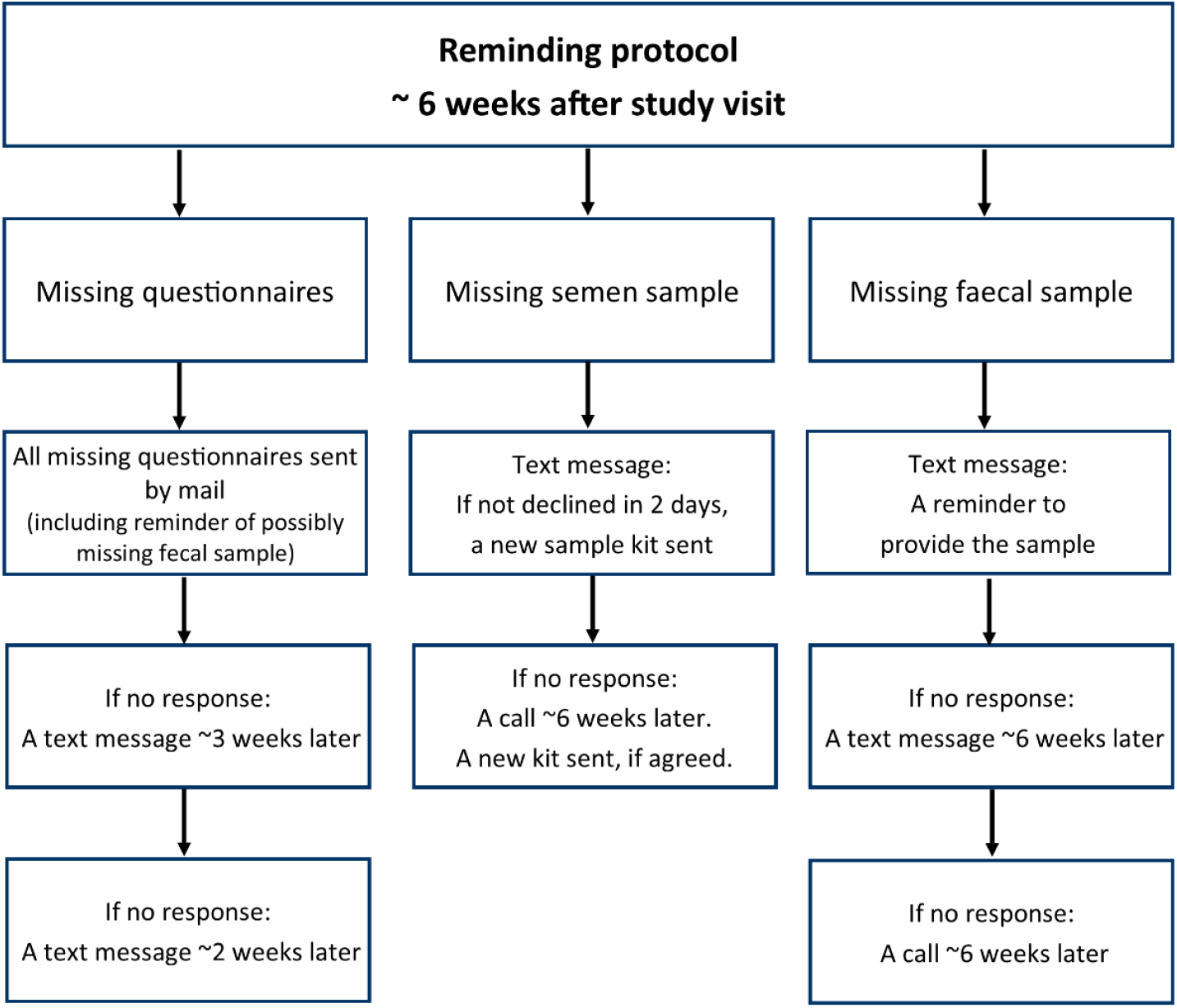
Illustration of the reminding protocol.

## What has it found? Key findings and publications

Table 2 presents the generation-specific proportion (%) and number (n) of provided data. In general, participants who attended the study visit were also active in completing the questionnaires. With regard to the samples, a blood sample was obtained from nearly all of those who attended the study visit. Semen samples were best provided by the G1 participants, followed by G2 and G0 participants. Data on anthropometrics, blood pressure, ultrasonographic measures, and cognitive function were obtained from nearly all study visit participants.

**Table 2.** Numbers (% of all invited / % of those who attended the study visit) of participants who provided data according to the generations.

|  | **G0** | **G1** | **G2<sup>a</sup>** | **Total** |
| --- | --- | --- | --- | --- |
|  | **n (%)** | **n (%)** | **n (%)** | **n (%)** |
| **Questionnaires<sup>b</sup>** |  |  |  |  |
| **Basic** | 2419 (61.4) | 2046 (63.6) | 2652 (46.6) | 7117 (55.4) |
|  | 2124 (99.0) | 1985 (96.2) | 2443 (96.1) | 6552 (97.0) |
| **Diet<sup>c</sup>** | 2242 (56.9) | 1829 (56.9) | 2166 (38.0) | 6237 (48.5) |
|  | 2040 (95.1) | 1797 (87.1) | 2078 (81.7) | 5915 (87.6) |
| **Psychosocial well-being** | 2309 (58.6) | 1796 (55.8) | 2202 (38.7) | 6307 (49.1) |
|  | 2096 (97.7) | 1765 (85.5) | 2114 (83.1) | 5975 (88.5) |
| **Samples** |  |  |  |  |
| **Blood** | 2134 | 2058 | 2390 | 6582 |
|  | (54.2 / 99.4) | (64.0 / 99.7) | (42.0 / 94.0) | (51.2 / 97.5) |
| **Semen<sup>d</sup>** | 289 | 612 | 388 | 1289 |
|  | (18.0 / 34.2) | (39.8 / 65.7) | (24.3 / 63.2) | (27.2 / 53.9) |
| **Hair** | 1676 | 1377 | 1815 | 4868 |
|  | (42.5 / 78.1) | (42.8 / 66.7) | (31.9 / 71.4) | (37.9 / 72.1) |
| **Faecal** | 1920 | 1476 | 1246 | 4642 |
|  | (48.7 / 89.5) | (45.9 / 71.5) | (21.9 / 49.0) | (36.1 / 68.7) |
| **Weight and height<sup>e</sup>** | 2138 | 2064 | 2537 | 6739 |
|  | (54.3 / 99.6) | (64.2 / 100.0) | (44.5 / 99.8) | (52.4 / 99.8) |
| **Blood pressure** | 2134 | 2057 | 2524 | 6715 |
|  | (54.2 / 99.4) | (63.9 / 99.7) | (44.3 / 99.3) | (52.2 / 99.4) |
| **Ultrasonography** |  |  |  |  |
| **Vascular<sup>f</sup>** | 2139 | 2062 | 2524 | 6725 |
|  | (54.3 / 99.7) | (64.1 / 99.9) | (44.3 / 99.3) | (52.3 / 99.6) |
| **Liver<sup>g</sup>** | 2139 | 2063 | 2525 | 6727 |
|  | (54.3 / 99.7) | (64.1 / 100) | (44.3 / 99.3) | (52.3 / 99.6) |
| **Cognitive function** | 2025 | 2030 | 2516 | 6571 |
|  | (51.4 / 94.4) | (63.1 / 98.4) | (44.2 / 98.9) | (51.1 / 97.3) |

<sup>a</sup>age-group specific data are given in the Supplement
<sup>b</sup>for questionnaires, the first numbers and proportions (%) are of all invited, and the second numbers and proportions (%) are of those who attended the study visit
<sup>c</sup>Food Frequency Questionnaire
<sup>d</sup>samples including sperm, the proportion of men who were aged $\geq 18$ years at the beginning of the study/reached the age by the study visit
<sup>e</sup>number/proportion of those with height and/or weight
<sup>f</sup>number/proportion of those with any vascular imaging data
<sup>g</sup>number/proportion of those with successfully obtained liver fat data

With respect to the key research aims of the MULTIEPIGEN project, 2020 (62.2%) G1 participants were exposed to paternal smoking and 1993 (66.0%) to parental psychosocial stress. Correspondingly, among the G2 participants, 1024 (46.8%) were exposed to paternal smoking and 1343 (66.0%) to paternal psychosocial stress (Table 3).

**Table 3.** Exposure to paternal smoking (n exposed / N data available) and psychosocial stress in early life in the generations 1 and 2.

| Exposure | Generation | Timing of the exposure |  |  |  |  |  |
| --- | --- | --- | --- | --- | --- | --- | --- |
|  |  | At conception (n/N) | During pregnancy (n/N) | Early childhood (n/N) | Childhood/adolescence (n/N) | Anytime before birth (n/N) | Anytime in childhood (n/N) |
| Smoking | G1 | 412/860 | 381/813 | 749/1509 | 1409/3141 | 768/1689 | 2020/3228 |
|  | G2 | 682/1650 | 653/1639 | 855/2003 | 775/1857 | 1670/2768 | 1024/2190 |
| Psychosocial stress | G1 | 98/827 | 80/794 | 731/3071 | 1520/3265 | 1383/3476 | 1993/3019 |
|  | G2 | 159/1511 | 170/1507 | 804/1890 | 877/1877 | 1887/1971 | 1343/2097 |

Early childhood: 0-6 years
Childhood/adolescence: 7-18 years

Semen samples were provided by 289, 612, and 388 male participants in generations G0, G1 and G2, respectively. Of these samples, 111, 418, and 294 had a successful RNA extraction (Supplementary Figure 1). These successfully extracted RNA samples include 22 G0-G1participant dyads, 55 G1-G2 dyads, 5 G0-G2 dyads, and one triad including all generations. Furthermore, the samples include 71 G2-G2 dyads (i.e. two brothers who have both given a sample), of which 14 are included in the G1-G2 dyads (i.e. their father has also given a sample) and one is included in the G0-G2 dyads (their grandfather has also given a sample). There are also 12 G2-G2-G2 triads (i.e. samples from three brothers), of which three are included in the G1-G2 dyads (their father has also given a sample). Additionally, a successful DNA extraction for methylation analysis was completed from 800 sperm samples. Of these, 149 were from G0 participants, 382 from G1 participants, and 269 from G2 participants. Taken together, successful extraction of both RNA and DNA was completed from 721 sperm samples. Of these, 100 were from G0 participants, 358 from G1 participants, and 263 from G2 participants.

Attrition analyses were conducted for the original cohort members (G1). Those who provided data in the latest follow-up were somewhat older and more often females than those who did not participate (Table 4). However, the participants and non-participants were similar in terms of baseline (year 1980) body mass index, systolic and diastolic blood pressure, serum total-, HDL-and LDL-cholesterol and triglycerides as well as the original study site.

**Table 4.**
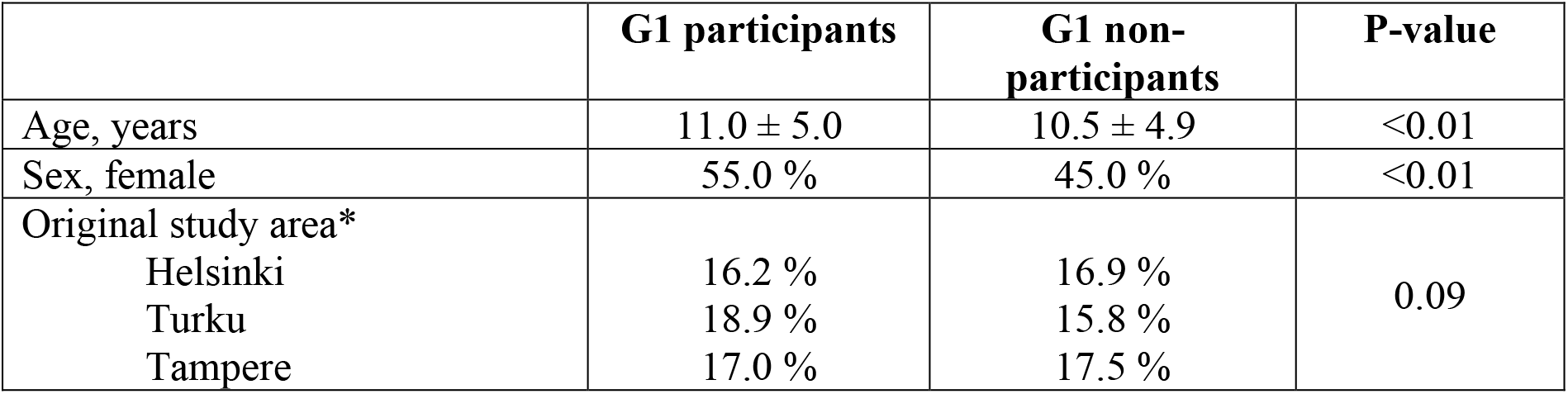
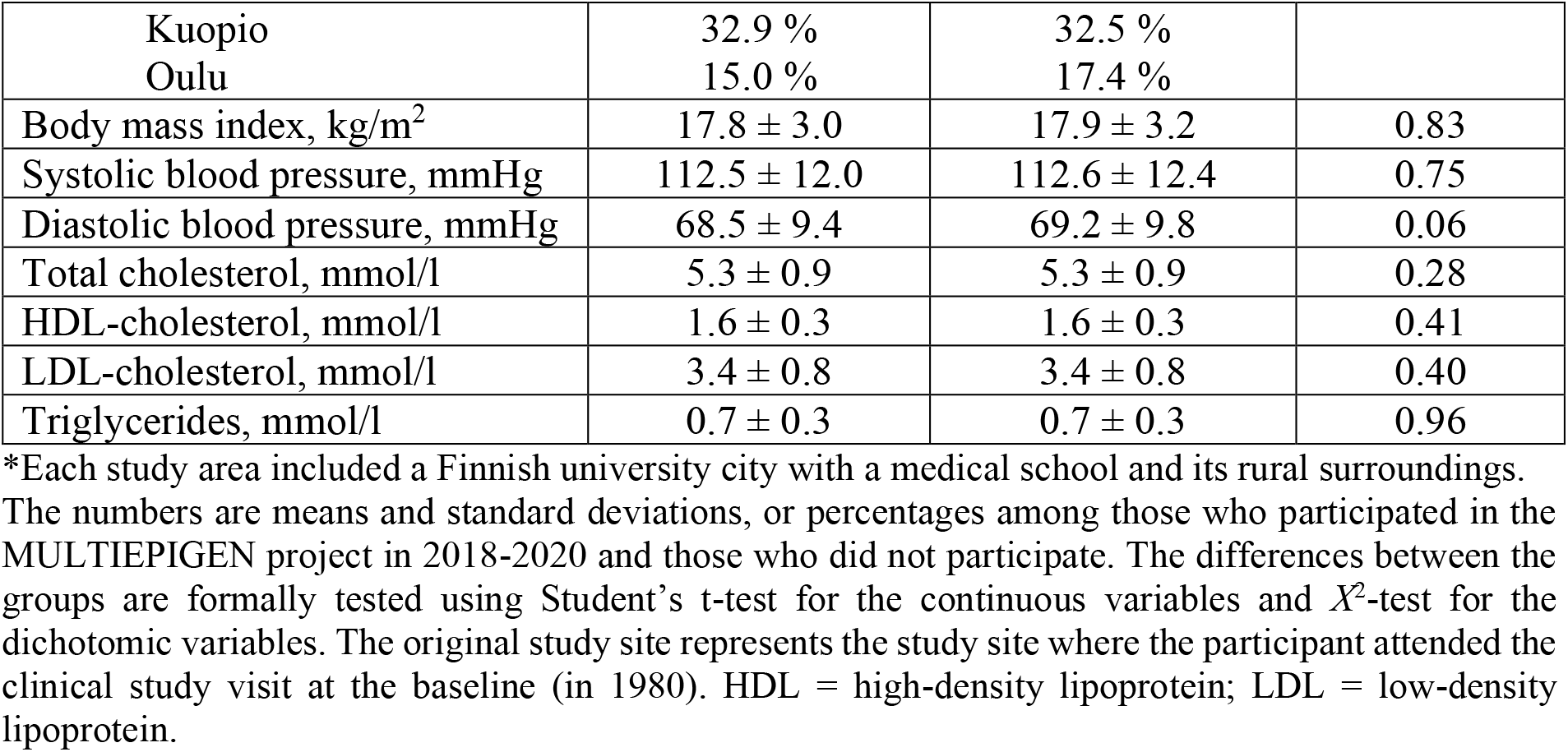
Baseline (year 1980) characteristics of the original G1 participants according to their participation in the latest follow-up study in 2018-20.

|  | G1 participants | G1 non-participants | P-value |
| --- | --- | --- | --- |
| Age, years | 11.0 ± 5.0 | 10.5 ± 4.9 | <0.01 |
| Sex, female | 55.0 % | 45.0 % | <0.01 |
| Original study area* |  |  |  |
| Helsinki | 16.2 % | 16.9 % | 0.09 |
| Turku | 18.9 % | 15.8 % |  |
| Tampere | 17.0 % | 17.5 % |  |
| Kuopio | 32.9 % | 32.5 % |  |
| Oulu | 15.0 % | 17.4 % |  |
| Body mass index, kg/m<sup>2</sup> | 17.8 ± 3.0 | 17.9 ± 3.2 | 0.83 |
| Systolic blood pressure, mmHg | 112.5 ± 12.0 | 112.6 ± 12.4 | 0.75 |
| Diastolic blood pressure, mmHg | 68.5 ± 9.4 | 69.2 ± 9.8 | 0.06 |
| Total cholesterol, mmol/l | 5.3 ± 0.9 | 5.3 ± 0.9 | 0.28 |
| HDL-cholesterol, mmol/l | 1.6 ± 0.3 | 1.6 ± 0.3 | 0.41 |
| LDL-cholesterol, mmol/l | 3.4 ± 0.8 | 3.4 ± 0.8 | 0.40 |
| Triglycerides, mmol/l | 0.7 ± 0.3 | 0.7 ± 0.3 | 0.96 |

\*Each study area included a Finnish university city with a medical school and its rural surroundings. The numbers are means and standard deviations, or percentages among those who participated in the MULTIEPIGEN project in 2018-2020 and those who did not participate. The differences between the groups are formally tested using Student's t-test for the continuous variables and $\chi^2$ -test for the dichotomic variables. The original study site represents the study site where the participant attended the clinical study visit at the baseline (in 1980). HDL = high-density lipoprotein; LDL = low-density lipoprotein.

## What are the main strengths and weaknesses?

The main strength of the multigenerational YFS is extensive number of well-phenotyped families including three generations. Additionally, the majority of the data collection methods were identical between the generations. Another strength is that it was built around the ongoing cohort. Thus, the multigenerational YFS benefits from the longitudinal study design covering 40 years, beginning from the childhood and being accompanied by regular follow-ups. Additionally, the collection of a diverse set of carefully measured phenotypes, lifestyle measures, and information on socioeconomic background provide the possibility to test and integrate at the population-level such hypotheses that have emerged from experimental studies. YFS is one of the most well-characterised life-course datasets globally with repeated and detailed measurements and sample collections. The YFS cohort offered several advantages serving as a base-population for a unique multigenerational epidemiologic study. Its existing biobank, containing serial serum sample collections from childhood to adulthood, enables the measurement of exposures to both intrinsic and extrinsic stressors. The study also benefits from linkage to the comprehensive national health registries maintained in Finland, such as morbidity, mortality, infant health and socioeconomic status (e.g. earnings and education). The multigenerational YFS therefore offers unique opportunities to study intergenerational transmission in humans in relation to several parental exposures and outcomes in their offspring.

Possible weaknesses include that the generalisability of the potential findings is limited to White Northern European population. Despite the intensive recruitment strategy, the number of participants remained lower than anticipated, especially regarding the number of semen sample donors. Additionally, in a multigenerational setting, it is challenging to form entire families including participants from all three generations. Finally, we acknowledge that our observational data cannot provide unequivocal proof of causality and that our longitudinal population-based study may be subject to different sources of bias (e.g. memory and selection bias, loss-to-follow-up, unmeasured confounding factors).

## Can I get hold of the data? Where can I find out more?

Due to the local legal restrictions concerning the distribution of all personal information, an open access to the data is not possible. Therefore, data sharing outside the study group requires a data-sharing agreement. Investigators can submit an expression of interest to the YFS Data Sharing Committee (youngfinnsstudy.utu.fi). After approval, de-identified datasets will be made available to the researcher. Data may be shared by direct transfer of data files via secured transfer systems or, especially in circumstances in which there is a risk of subject identification or when security or other legal restrictions apply to the data, a data enclave may be provided as a physical location where the user can access shared data under controlled conditions.

## Supporting information

Statistical analysis plan

## Data Availability

Due to the local legal restrictions concerning the distribution of all personal information, allowance of open access to the YFS data is not possible. Therefore, data sharing outside the study group requires a data-sharing agreement. Investigators can submit an expression of interest to the YFS Steering Group / Data Sharing Committee (PI of the YFS, Prof. Olli Raitakari).

## Ethics approval

Ethical committees of the Hospital District of Southwest Finland and the European Research Council have approved the study. All individuals and/or legal guardians have signed a written informed consent to take part in the study. They had the right to refuse any part of the study protocol or discontinue at any time without the need to give any explanation.

## Author contributions

Study conception and design:, KP, SR, ME, MH, AJ, MJ, MK, NKa, IL, TL, J-AM, LPR, TR, JT, JV, OR

Carried out data collection: KP, SR, MB, ME, JH, NH, EJ, MJ, HK, JK, MK, NKo, IL, SM, LPR, SS, LT, PT, THT, OR,

Designed the analytical strategy: KA, NKa, JN

Supervised laboratory analyses: MB, B-ML, TL, JM, ER, PR

Performed data preparation: KP, SR, MB, ME, JH, IL, L-PL, PPM, LPR, ER, SS, THT Carried out data analysis: MB, JH, NKa, JSK, IL, L-PL, CM, PPM, SS,

Drafted the manuscript: KP, SR, ME, JH, NKa, JM, SM, J-AM, LPR, SS, THT, OR

All authors critically revised the manuscript for intellectual content. All authors read and approved the final manuscript of the paper. KP, SR and OR are guarantors of the paper.

## Funding

The Young Finns Study has been financially supported by the Academy of Finland: grants 356405, 322098, 286284, 134309 (Eye), 126925, 121584, 124282, 129378 (Salve), 117797 (Gendi), and 141071 (Skidi); the Social Insurance Institution of Finland; Competitive State Research Financing of the Expert Responsibility area of Kuopio, Tampere and Turku University Hospitals (grant X51001); Juho Vainio Foundation; Paavo Nurmi Foundation; Finnish Foundation for Cardiovascular Research; Finnish Cultural Foundation; The Sigrid Juselius Foundation; Tampere Tuberculosis Foundation; Emil Aaltonen Foundation; Yrjö Jahnsson Foundation; Signe and Ane Gyllenberg Foundation; Diabetes Research Foundation of Finnish Diabetes Association; EU Horizon 2020 (grant 755320 for TAXINOMISIS and grant 848146 for To Aition); European Research Council (grant 742927 for MULTIEPIGEN project); Tampere University Hospital Supporting Foundation; Finnish Society of Clinical Chemistry; the Cancer Foundation Finland; pBETTER4U_EU (Preventing obesity through Biologically and bEhaviorally Tailored inTERventions for you; project number: 101080117); CVDLink (EU grant nro. 101137278) and the Jane and Aatos Erkko Foundation; Research Committee of the Kuopio University Hospital Catchment Area (State Research Funding, 5031364); Research Council of Finland’s Flagship InFLAMES (funding decision numbers 337530 and 357910). Pashupati P. Mishra was supported by the Academy of Finland (Grant number: 349708) and Emma Raitoharju (grants: 330809, 338395).

## Acknowledgements

The YFS families have made this study possible — the authors thank them for their time, efforts, and commitment to the YFS throughout the years. Faecal metagenome sequencing and **s**perm DNA methylation sequencing was performed in Finnish Functional Genomics Centre Facility supported by University of Turku, Åbo Akademi University and Biocenter Finland.

## Conflict of interest

The authors have no conflicts of interest.

## Notes

### Competing Interest Statement

The authors have declared no competing interest.

## References

1. Raitakari OT, Juonala M, Rönnemaa T, Keltikangas-Järvinen L, Räsänen L, Pietikäinen M, et al. Cohort profile: The Cardiovascular Risk in Young Finns Study. Int J Epidemiol. 2008;37:1220–6.

2. Åkerblom HK, Viikari J, Uhari M, Räsänen T, Byckling T, Louhivuori K et al. Atherosclerosis precursors in Finnish Children and Adolescents. 1. General description of the cross-sectional study of 1980, and an account of the children’s and families state of health. Acta Paediatr Scand Suppl 1985;318:49–63.

3. Juonala M, Magnussen CG, Berenson GS, Venn A, Burns TL, Sabin MA, et al. Childhood Adiposity, Adult Adiposity, and Cardiovascular Risk Factors. N Engl J Med. 2011;365:1876–1885.

4. Jacobs DR, Woo JG, Sinaiko AR, Daniels SR, Ikonen J, Juonala M, et al. Childhood Cardiovascular Risk Factors and Adult Cardiovascular Events. N Engl J Med. 2022;386:1877–88.

5. Wu F, Jacobs DR, Daniels SR, Kähönen M, Woo JG, Sinaiko AR, et al. Non–High-Density Lipoprotein Cholesterol Levels From Childhood to Adulthood and Cardiovascular Disease Events. JAMA. 2024;331:1834.

6. Phelps NH, Singleton RK, Zhou B, Heap RA, Mishra A, Bennett JE, et al. Worldwide trends in underweight and obesity from 1990 to 2022: a pooled analysis of 3663 population-representative studies with 222 million children, adolescents, and adults. Lancet. 2024;403:1027–50.

7. Karjalainen MK, Karthikeyan S, Oliver-Williams C, Sliz E, Allara E, Fung WT, et al. Genome-wide characterization of circulating metabolic biomarkers. Nature. 2024;628:130–8.

8. Pembrey M, Saffery R, Bygren LO, Network in Epigenetic Epidemiology. Human transgenerational responses to early-life experience: potential impact on development, health and biomedical research. J Med Genet. 2014;51:563–72.

9. Chen Q, Yan M, Cao Z, Li X, Zhang Y, Shi J, et al. Sperm tsRNAs contribute to intergenerational inheritance of an acquired metabolic disorder. Science. 2016;351:397– 400.

