## Supplemental material for "Cohort Profile Update: Expanding the Cardiovascular Risk in Young Finns Study into a multigenerational cohort"

### Supplementary Material

This document describes the data collections organised in the multigenerational Young Finns Study. One key research theme relates to epigenetic inheritance (MULTIEPIGEN project). The MULTIEPIGEN examines whether paternal exposures to certain stressors are linked with offspring phenotypes mediated by epigenetic markers in the sperm. The pre-specified paternal and grandpaternal exposures of interest include smoking, psychosocial adversities, adiposity and serum levels of persistent organic pollutants. The pre-specified offspring outcomes include cardio-metabolic phenotypes, such as components of the metabolic syndrome (BMI, waist circumference, blood pressure, serum lipids and apolipoproteins, serum insulin and glucose, fatty liver), markers of vasculopathy indicative of atherosclerosis (carotid artery intima-media thickness and elasticity, and carotid artery plaques), markers of cognitive function (episodic memory and associative learning, working memory, executive function, reaction and movement time, sustained attention), and markers of psychological wellbeing (quality of life, depression, anxiety, and quality of sleep). The pre-specified mediators include sperm non-coding RNA profile and sperm DNA methylation markers.

In the method descriptions below, generation 0 (G0) refers to the first generation, i.e. the parents of the original Young Finns participants, generation 1 (G1) refers to the original Young Finns participants, and generation 2 (G2) refers to the offspring of the original Young Finns participants.

#### Pre-specified paternal exposures

*Paternal smoking exposure*

Longitudinal data on the smoking behaviour of the G1 participants have been prospectively acquired using questionnaires collected in multiple follow-up studies conducted during their childhood, adolescence and adulthood. Prospective longitudinal data on the smoking behaviour of the G0 participants have been acquired using questionnaires collected during the childhood and adolescence of the G1 participants.

In the MULTIEPIGEN project, we used a retrospective questionnaire to assess parental smoking at different developmental stages of the offspring. The G0 participants were asked to fill in the questionnaire regarding each of their G1 offspring, whereas the G1 participants were asked to fill in the questionnaire regarding each of their G2 offspring. Specifically, in the retrospective questionnaire, we asked about the frequency and intensity of smoking at four time periods: at conception of the offspring; during pregnancy; when the offspring was under 7 years of age; and when the offspring was aged between 7 and 18 years.

The retrospective data on parental smoking were transformed into a longitudinal trajectory on a yearly basis corresponding to the offspring participants' age. Smoking status at the time of conception was assigned on the year before the birth year of the participant. Smoking status at the time of pregnancy was assigned on the birth year of the participants. Smoking status before and after the offspring was 7 years of age was assigned for the years when the participant was between 1 and 6 years of age and between 7 and 18 years of age, respectively. These trajectories were built for the questionnaires of each offspring separately and thus parents with multiple offspring had multiple longitudinal, possibly overlapping trajectories. After building the trajectories on based on the retrospective questionnaire data, information based on the retrospective reporting was combined with the prospective information.

Any discrepancy between the prospective and retrospective smoking status was solved on a case-by-case level, utilizing all smoking information available for the individual, e.g. information on smoking cessation. A random sample of case-by-case solutions was reviewed by another coder to ensure the validity of the solutions. The solutions were blinded for any other variables except for those that describe the smoking patterns of the parent throughout the life-course.

The final longitudinal smoking trajectories were summarised into measures of smoking at different stages of offspring development. The variables include the initial developmental periods of interest (i.e., at the time of conception, during pregnancy, in early childhood, during school age) as well as more comprehensive summary variables (parental life-course smoking at any time before conception, parental smoking at any time at offspring childhood). The new summarised variables based on both prospective and retrospective data are available from a larger population than those based on only prospective or retrospective information, and may be considered more valid as the discrepancies between the two sources of information were inspected.

*Paternal exposure to stressful life events (psychosocial adversities)*
Longitudinal data on stressful events of the G1 participants have been prospectively collected with questionnaires in the years 1986, 1989, 1992, 2001, 2007 and 2018. Stressful events included meaningful life experiences, such as death of a close friend, family member or relative, divorce or separation, illness, long-term unemployment or other important changes in life (moving residence, moving school, switching position). In addition, catastrophic life-experiences were inquired in 2018, these include encountered (mental or physical) violence or a natural disaster. All the listed events were regarded as exposure events for the offspring with one exception: the G2 participants' own birth, which was sometimes reported as a life experience by G1 participants in their 2007 questionnaire, was excluded as an exposure event for the G2 participant. Furthermore, few stressful events for the G0 participants have been collected with questionnaires in the years 1980 and 1983.

To construct three-generational exposure data, we used a retrospective questionnaire to assess stressful events at the time of the conception of the offspring, and during pregnancy. The G0 participants were asked to fill in the questionnaire regarding each of their G1 offspring, whereas the G1 participants were asked to fill in the questionnaire regarding each of their G2 offspring. The questions were about the worsening of their family's economic situation; involuntary termination of unemployment; divorce; own or family member's serious illness; death of their spouse, own child or a close friend. There were also open-ended questions for other stressful life events related to living (e.g. moving or building house), problems with significant other and health, money, work or study problems.

After the events were assigned to a year-by-year level, we then combined the G0 participants' events to their G1 offspring data and the G1 participants' events to their G2 offspring data. Subsequently, we evaluated how many, if any, stressful events the G1 and G2 participants had been exposed to by their parent during the periods of conception (one year before birth), pregnancy (birth year), early childhood (ages 0-6 years) and childhood (ages 7-18 years).

*Paternal adiposity*
Paternal adiposity was assessed at the 2018 clinic visit by measuring height, weight and waist circumference and calculating body mass index ($kg/m^2$). In addition to the continuous measures, the measures were categorised. Males with waist circumference >102 cm were categorised as having central obesity. Dichotomous variables describing overweight and obesity were calculated using the cut-offs 25 and 30 for BMI, respectively.

In addition to these measures, prospective data were used specifically to assess the epigenome-wide as well as the inter- and transgenerational effects of adiposity earlier in life.

For G1 fathers, we calculated the long-term burden of BMI and adiposity by modelling the subject-specific longitudinal measurements using mixed-effect regression splines as reported previously.(1,2) Briefly, the spline fits provide curves for each individual at each age point, and the cumulative burden can be calculated by deriving an area under the curve (AUC) within the desired age interval. We here used the AUCs for age periods 6-12, 12-18 and 18-24. The BMI splines were based on longitudinal BMI measures while the adiposity splines were based on sum of triceps, subscapular and biceps skinfold measures in childhood and on waist measurements in adulthood, both standardised to mean 100 and standard deviation 15. In addition, we utilised the longitudinal data and spline estimates to build a variable that describes the paternal BMI nearest to the time of offspring conception.

For G0 fathers, we used their prospective BMI measures, reported at the beginning of the study (1980) or, if that was missing, in later questionnaires during the childhood of the G1 participant.

*Paternal exposure to environmental toxicants*
Altogether 22 persistent organic pollutants (POP) were analysed from the 1980 baseline and 2001 follow-up serum samples in G1. The POPs included 10 PCB congeners (74, 99, 118, 138, 153, 156, 170, 180, 183 and 187), 9 OCPs (PeCB, HCB, α-HCH, β-HCH, γ-HCH, oxy-chlordane, trans-nonachlor, p,p'-DDT and p,p'-DDE), and 3 PBDEs (47, 99 and 153). The method consisted of liquid-liquid extraction, silica column clean-up and analysis by Agilent 7010 gas chromatograph triple quadrupole mass spectrometer (GC-MS/MS). In addition, 13 PFAS compounds (PFHxA, PFHpA, PFOA, PFNA, PFDA, PFUnA, PFDoA, PFTrA, PFTeA, PFHxS, PFHpS, PFOS, PFDS) were analysed with a method that consisted of liquid-liquid extraction and analysis by Thermo Scientific UltiMate 3000 Rapid Separation LC system connected to Thermo Finnigan TSQ Quantum Discovery MAX triple quadrupole mass spectrometer (LC-MS/MS).

POPs were analysed in batches of 36 samples and PFAS in batches of about 70 samples, both in two sets. The first set of samples was analysed in 2016 and it included 1301 samples from baseline year 1980 and 1259 samples from the follow-up year 2001. The participants were chosen based on the availability of some key measures in both 1980 and 2001 (BMI, lipids, blood pressure, birth weight, SES; GWAS, dietary measures, parental smoking). This sample was also enriched with participants with type II diabetes by including all participants who had been diagnosed with type II diabetes by 2016. The second set of samples included all remaining eligible serum samples in 1980 (n=1129) and 2001 (n=972) as well as serum samples obtained in 1992 from 197 male study subjects who had given semen samples in the MULTIEPIGEN study. The analysis of the POPs has been finished from the second set and 13 PFAS compounds are currently being analysed.

The values that were below the detection limit (Supplementary Table 1) were set to the mid-point between zero and the detection limit. For example, when the detection limit was 5, the values below the detection limit were set to 2.5.

**Supplementary Table 1.** Detection limits of the POP and PFAS compounds.

| Compound | Detection limit |
|---|---|
| PCB74 | 5 pg/ml |
| PCB99 | 5 pg/ml |
| PCB118 | 5 pg/ml |
| PCB153 | 5 pg/ml |
| PCB138 | 5 pg/ml |
| PCB156 | 5 pg/ml |
| PCB170 | 5 pg/ml |
| PCB180 | 5 pg/ml |
| PCB183 | 5 pg/ml |
| PCB187 | 5 pg/ml |
| PeCB | 10 pg/ml |
| HCB | 10 pg/ml |
| α-HCH | 20 pg/ml$^{+}$ |
| β-HCH | 15 pg/ml |
| γ-HCH | 20 pg/ml |
| oxy-chlordane | 25 pg/ml |
| trans-nonachlor | 5 pg/ml |
| p,p'-DDT | 15 pg/ml |
| p,p'-DDE | 40 pg/ml |
| BDE47 | 15 pg/ml |
| BDE99 | 15 pg/ml |
| BDE153 | 15 pg/ml |
| PFHxA | 0.3 ng/ml |
| PFHpA | 0.2 ng/ml |
| PFOA | 0.2 ng/ml |
| PFNA | 0.2 ng/ml |
| PFDA | 0.2 ng/ml |
| PFUnA | 0.2 ng/ml |
| PFDoA | 0.3 ng/ml$^{*+}$ |
| PFTrA | 0.5 ng/ml$^{*+}$ |
| PFTeA | 0.5 ng/ml$^{*+}$ |
| PFHxS | 0.2 ng/ml |
| PFHpS | 0.2 ng/ml |
| PFOS | 0.2 ng/ml |
| PFDS | 0.5 ng/ml$^{*}$ |

$^{+}$all participants were below the detection limit in 2001
$^{*}$all participants were below the detection limit in 1980

The compounds within the same chemical group (e.g. PCBs) tend to be highly correlated with each other. In our analyses, the primary toxicant exposures of interest are summary measures for each group of toxicant compounds (PFAS, PCB, BDE, Organochlorine). Before summarization, each raw congener variable was first log-transformed to correct the skewness in their distribution. Then, each variable was standardised to mean 0 and variance 1. The summary measures were then calculated as the sum of the compounds within each group and year. As an additional analysis, each compound is studied separately.

**Pre-specified offspring outcomes**

The pre-specified offspring outcomes include cardio-metabolic phenotypes, such as components of the metabolic syndrome (BMI, waist circumference, blood pressure, serum lipids, serum insulin, fatty liver), markers of vasculopathy indicative of atherosclerosis (carotid artery intima-media thickness and elasticity, and carotid artery plaques), markers of cognitive function (episodic memory and associative learning, working memory, executive function, reaction and movement time, sustained attention), and markers of psychological wellbeing (quality of life, depression, anxiety, and quality of sleep).

We describe below the measurement protocols related to these key offspring outcomes. Of note, while these measures are used as offspring outcomes, they were measured in a similar manner across all generations, unless specified otherwise. When necessary and possible, the paternal levels of these phenotypes are adjusted for in the inter- and transgenerational models.

*Offspring cardiometabolic outcomes*

*Anthropometrics* - Height, weight, and waist circumference were measured at the clinic visit, and body mass index (kg/$m^2$) was calculated. For BMI, dichotomous variables describing overweight and obesity were calculated using the cut-offs according to Cole criteria.(3) Central adiposity was defined based on waist circumference. The cut-off was 88 centimetres for adult (aged 18 or older) females and 102 centimetres for adult males. For children and adolescents aged less than 18 years, we calculated the relationship of waist circumference and height. If this relationship was larger than 0.5, the participants were categorised as having central adiposity.

*Blood pressure* - From all participants, blood pressure was measured similarly in a sitting position primarily from the right arm brachial artery (if no possible, from the left arm) with automatic Omron HBP-1300 blood pressure measurement device (Omron Corporation, Kyoto, Japan). The cuff size was determined after measuring forearm circumference 10 cm above elbow joint. During the measurement, the arm rested so that the elbow joint was set at the heart level and feet rested separately on the floor. The participant was asked not to move or talk during the measurement. After 3 minutes of rest, blood pressure was measured three times at 1-minute intervals. If the difference between the second and the third measurement was ≥15 mmHg in adults, fourth measurement was done. From the third and fourth measurement, the blood pressure value closer to second measurement was recorded. The mean of three measurements was calculated.

In addition, an automated oscillometric device (Mobil-O-Graph; I.E.M., Stolberg, Germany) was used to determine right arm brachial and aortic blood pressure, heart rate, pulse wave velocity (PWV), stroke index (SI; stroke volume/body surface area, ml/$m^2$), cardiac index (CI; cardiac output/body surface area, l/min/$m^2$) and systemic vascular resistance index (SVRI; systemic vascular resistance/body surface area, dyn·s/cm5/$m^2$) in supine posture (all three generations) and in standing posture after 3 minutes of standing (G1). The cuff size was determined after measuring forearm circumference 10 cm above elbow joint.

*Lipids, glucose and insulin* - Venous blood samples were taken after a minimum of a 4-hour fast and stored at -70°C until analysis. Lipids and glucose were determined from serum samples with enzymatic assays using the following system reagents; HDL-Cholesterol Plus, Cholesterol, Triglycerides and Glucose (HK) (all from Thermo Fisher Scientific, Finland) on an Indiko Plus analyzer (Thermo Fisher Scientific, Finland). Insulin was determined from serum samples by a chemiluminescent microparticle immunoassay using the LIAISON® Insulin system reagent (DiaSorin S.p.A., Italy) on a Liaison XL immunoassay analyzer (DiaSorin Deutschland GmbH, Germany). LDL-cholesterol was estimated with Friedewald's formula (for participants with triglycerides concentration <4.5mmol/l).(4)

*Carotid artery structure and function* - Carotid ultrasound studies were performed using GE Logiq S8 (GE Vingmend Ultrasound A/S, Horten, Norway) ultrasound mainframes with a ML6-15-D matrix linear transducer. For children with shorter necks, 9-L linear transducer was also used. Carotid ultrasound studies were performed according to standardised protocols by sonographers and trained ultrasound technicians to assess preclinical vascular changes related to atherosclerosis; intima-media thickness (IMT) of the left common carotid artery, carotid bifurcation, and internal carotid artery, left common carotid artery elasticity, longitudinal movement of the left common carotid arterial wall, and identification of plaques within the left and right common carotid artery, carotid bifurcation, and internal carotid artery. Participants were examined in supine position with their necks extended. During the ultrasonography, a continuous electrocardiogram was recorded.

*Carotid intima-media thickness* - A frequency of 15 MHz was used to assess the left common carotid IMT (cIMT). The image was focused on the distal part of the common carotid artery so that both the intima of the near and far walls were visible. To achieve a maximum frame rate, a single focus point was used. The zoom function was used to capture an image 35 mm wide and 25 mm high, including the distal part of the common carotid artery and the beginning of carotid bifurcation. A 5-second moving clip was captured and stored for subsequent offline cIMT analysis. For the assessment of carotid bifurcation IMT (bIMT), the transducer was moved distally along the carotid artery to include the distal part of the common carotid artery and carotid bulb in the image. The frequency could be lowered, ensuring sufficient tissue penetration to optimise image quality. If possible, the intima of both near and far walls of the carotid bifurcation were made visible, and a single focus point was used to focus the far wall of the carotid bifurcation. Then, a 5-second moving clip was captured and stored for subsequent offline bIMT analysis. For the assessment of internal carotid IMT (iIMT), the transducer was moved distally to include the distal part of the carotid bulb and internal carotid artery in the image. Pulsed wave and colour Doppler was utilised to differentiate between the internal and external carotid arteries. The internal carotid artery was characterised by a lower systolic velocity and a higher diastolic velocity compared to the external carotid artery. Additionally, colour Doppler was used in identifying typical side branches of the external carotid artery. The image was then focused on the internal carotid artery by using a single focus point. If possible, the intima of both the near and far walls of the internal carotid artery were made visible. Then, a 5-second moving clip was captured and stored for subsequent offline iIMT analysis. IMT measurements were obtained with a semi-automated TOMTEC AutoIMT (version TTA2.41.00, TOMTEC Imaging Systems GmbH, Unterschleissheim, Germany) by two readers who were blinded to participant details. For cIMT measurements, the highest quality end-diastolic frame (coincident with the R wave on a continuously recorded electrocardiogram) was selected from the 5-second clip. On the selected frame, a box approximately 10 mm wide (region of interest) was delineated with TOMTEC AutoIMT, positioned 5 to 15 mm proximal to the carotid bifurcation. Then, the program automatically identified the lumen-intima and media-adventitia interfaces of the far wall, drew lines at these interfaces, and automatically calculated the mean cIMT for the specified region of interest. Carotid bIMT and iIMT were measured with a similar semi-automated technique. Carotid bIMT was measured approximately from a 3-mm segment starting from the proximal part of the carotid bifurcation and iIMT approximately from a 4-mm segment ~10 mm distal to the flow divider. Automated IMT measurements were approved by the reader. If the automatic detection of arterial interfaces was not satisfactory, the reader performed four manual IMT measurements from the defined region of interest. For cIMT, manual measurements were made approximately 10 mm proximal to the carotid bifurcation.(5) The mean of these four manual measurements was then used in the statistical analysis. IMT measurements were performed from the plaque-free area.

*Carotid artery plaque* - During ultrasonography, both the left and right carotid arteries were scanned to detect carotid artery plaques. These plaques were defined as focal structures protruding into the arterial lumen of at least 0.5 mm or 50% of the surrounding IMT value or demonstrating a thickness of 1.5 mm as measured from the media-adventitia interface to the intima-lumen interface.(6) When a carotid plaque was suspected, it was scanned from the longitudinal view to ensure accurate visualization of the plaque-adventitia and plaque-lumen interfaces. A 5-second moving clip was then captured and stored for subsequent offline plaque analysis. For all participants, moving cross-sectional scans of both the left and right carotid arteries were taken, starting from the common carotid artery, and extending to the point where the internal and external carotid arteries had branched. These scans were stored for offline plaque analysis. Carotid artery plaque measures were performed by two experienced readers blinded to participant's details' These plaque measurements were obtained using imaging software packages (TOMTEC and ComPACS Viewer). First, ultrasound images were scanned to confirm plaque diagnostics according to plaque definition. The plaque area was then measured for each plaque from the highest-quality longitudinal frame. Manual tracing of plaque lumen-intima and media-adventitia boundaries was performed around the perimeter, and the area within these boundaries was automatically measured. The maximum thickness of the plaque was measured manually from the same frame, representing the thickest distance from the media-adventitia interface to the lumen-intima interface. For the determination of the maximum thickness of the thickest plaque in the carotid artery tree including the common carotid artery, carotid bifurcation, and internal carotid artery, a cross-sectional view was employed. The thickest plaque was manually measured from the media-adventitia interface to the lumen-intima interface in the cross-sectional view. The number of carotid plaques in each carotid segment was derived from the longitudinal view. In the G0 population, where plaque prevalence was higher, cross-sectional frames were used to calculate the plaque count.

*Carotid artery elasticity* - Common carotid artery elasticity was assessed by using M-mode frame positioned approximately 10 mm proximal to the carotid bifurcation. A captured M-mode image displayed the near and far wall intima and spanning a minimum of two cardiac cycles on a continuously recorded electrocardiogram. Then, the scan was stored for subsequent off-line elasticity analysis. Measurements of the common carotid artery diastolic and systolic diameters were taken as the distance from the near and far wall intima. These measurements were obtained from two cardiac cycles, and the mean of these measures was used to define carotid artery elasticity measures. Ultrasound and concomitant brachial blood pressure measurements were used to calculate the three indices of arterial elasticity: 1) carotid distensibility ([Ds-Dd]/Dd)/(Ps-Pd); 2) Young's elastic modulus ([Ps-Pd]xDd)/([Ds-Dd]/IMT);

and 3) stiffness index ln(Ps/Pd)/([Ds-Dd]/Dd). Dd is the diastolic diameter, Ds is the systolic diameter, Ps is systolic blood pressure, Pd is diastolic blood pressure, and IMT is common carotid artery intima-media thickness.

*Hepatic steatosis* - To assess liver fat content, hepatic ultrasound imaging with a validated protocol was performed. Logiq S8 (GE Healthcare, Chicago, IL, USA) ultrasound device with a 1.5-6.0 MHz convex C1-6 transducer was used. Estimation of hepatic steatosis was based on four or five of the following criteria: liver-to-kidney contrast, parenchymal brightness, deep beam attenuation, bright vessel walls, and visibility of the neck of the gallbladder. Both at baseline and at follow-up, the same trained sonographer evaluated hepatic steatosis visually from the ultrasound images according to the aforementioned criteria and classified the participants to have a fatty liver or a normal liver. The sonographer was masked to participants´ clinical characteristics.

*Cognitive function outcomes*
Cognitive testing was performed using a computer-based cognitive test battery (CANTAB®). The CANTAB® is a computerised, predominantly non-linguistic and culturally neutral test focusing on a wide range of cognitive domains. The test is performed using a validated touch-screen computer system. The full test battery includes several individual tests from which, a suitable test battery for each particular study may be selected. The test battery selected for the extended YFS field study included five separate tests that are sensitive to aging and capture variation even within the cognitively healthy cohort. The selected tests measure four cognitive domains: 1) visual and episodic memory and visuospatial associative learning, 2) short-term and spatial working memory and problem-solving, 3) reaction and movement speed, and 4) visual processing, recognition and sustained attention. The test battery was identical for all participants aged 7 years and older, while a partly modified and shorter version of the same test battery was selected for the children aged 3-6 years. A study nurse administered the test to all participants and ensured that there was no misunderstanding related to performing the test. Voice-over instructions were provided by the CANTAB® software to the older children, adolescents and adults. For the small children, the instructions were given by a study nurse. This procedure was selected to guarantee that the small children would understand the test instructions properly, and thus to ensure reliable data.

First, the participants conducted a *Motor Screening (MOT) test* measuring psychomotor speed and accuracy. In the YFS cognitive testing protocol, the MOT test was considered as a training procedure which introduced the test equipment for the participants. Simultaneously, the MOT test was used as a screening tool to point out any difficulties in vision, movement, comprehension or ability to follow the test instructions. During the MOT test, a series of red crosses were shown in different locations on the screen, and the participants were advised to touch, as quickly as possible, the center of the cross every time it appeared. The MOT test was identical for all participants regardless of age. After the MOT test, four separate tests each measuring a specific cognitive domain were conducted.

*Paired Associates Learning (PAL) test* measured visual and episodic memory and visuospatial associative learning containing aspects of both delayed response procedure and conditional learning. For the participants aged 7 years and older, either 2, 4, 6, 8, or 12 patterns were displayed sequentially in boxes placed on the screen during the PAL test. After that, the patterns were presented in the center of the screen, and the participants were supposed to point to the box in which the particular pattern was previously seen. The test moves on to the next stage if all the patterns were placed in right boxes. In case of an incorrect response, all the patterns were re-displayed in their original locations and another recall phase was followed. The test terminated if the patterns were still incorrectly placed after 4 presentation and recall phases. For the participants aged 3-6 years, the test was identical to the test conducted for older participants except that the 12-pattern stage was excluded.

*Spatial Working Memory (SWM) test* was used to measure the ability to retain spatial information and to manipulate items stored in the working memory, problem-solving as well as the ability to conduct a self-organised search strategy. During the SWM test, the participants aged 3 years and older were presented with either 3, 4, 6, 8, or 12 randomly distributed coloured boxes on the screen. After that, the participants were supposed to search for tokens hidden in the boxes. When a token was found it was supposed to be moved to fill an empty panel on the right-hand side of the screen. Once the token had been moved from the box, the participant had to recall that the computer would never hide a new token in a box that previously contained one; therefore, the participants were not supposed to revisit the same boxes again.

*Reaction time (RTI) test* assessed speed of response and movement on a task where the stimulus was unpredictable (five-choice location task). In the RTI test administered for the participants aged 7 years and older, five large circles were presented on the screen. The participant was supposed to press down a touchscreen button at the bottom of the screen and wait until a small yellow spot appeared in any of the five large circles. When the yellow spot appeared, the participant was supposed to touch the yellow spot as soon as possible with the same hand that was pressing the touchscreen button. The RTI test administered for the participants aged 3-6 years included an identical five-choice task to the test for older participants. However, in the beginning of the RTI test the small children performed also a simple-choice stage, where only one large circle was presented on the screen and the target appeared always in this circle.

*Rapid visual information (RVP) test* was used to assess visual processing, recognition and sustained attention. In this test, the participants aged 7 years and older were presented with three-number sequences (3-5-7, 2-4-6 and 4-6-8) next to a large box where numbers 1-9 appeared in a random order at a rate of 100 numbers per minute. Whenever any of the particular sequences were presented, the participant was supposed to press a touchscreen button. Altogether nine target sequences were presented in every 100-seconds interval during a six-minutes assessment phase. During the practice phase, the participant was given visual cues (*i.e.* coloured or underlined numbers) to help to recognise the particular sequence. At the assessment phase, the cues were no longer presented. For the participants aged 3-6 years, the test included only one number sequence (3-5-7).

Each of the CANTAB® tests produced several variables. We used Flury's common principal component analysis (7) to derive the principal component scores for (i) across all domains / whole CANTAB test battery and (ii) separately for each measured cognitive domain / each of the four separate subtests. The main idea of this method is to conduct a principal component analysis for a data set arranged in multiple groups. It allows the groups to have different means, variances, and correlations but assumes that the principal components are the same in those groups. As the cognitive function may differ between generations and participants of different ages, we defined six groups as the input for the analysis as: G0, G1, G2 (aged 25-40 years), G2 (18-24 years), G2 (13-17 years), and G2 (7-12 years). The G2 generation (children of the original YFS participants) was divided into four age windows, as we wanted to allow/recognize the possible variation of cognitive function within different developmental phases. We standardised the variables of cognitive function before the analysis and subsequently, winsorised few outlying values (>10 SDs from the mean) to +/-10 to control any disproportionate influence. The common principal component analysis was implemented with the multigroup package in R (version 4.1.3).

For the purposes of statistical analyses, the first principal component (PC) within each cognitive domain was normalised for age and sex through the utilization of unexplained residuals. First, the mean behaviour of each PC in each cognitive domain was modelled for both sexes separately by fitting a smoothing spline to the relationship between the principal component and age. The smoothing parameters were chosen using leave-one-out cross-validation. Spline models were employed to accommodate the non-linear nature of cognitive changes across ages. Subsequently, the unexplained residual was computed as the difference between an individual participant's observation and the age-specific spline-derived value.

**Psychosocial outcomes**
The main psychological wellbeing outcomes were depressive symptoms, anxiety, and quality of life. These dimensions were chosen to reflect mental health deficits (depressive and anxiety symptoms) and positive mental health resources (questionnaires on quality of life). Psychosocial factors were self-reported by adult participants (aged ≥ 18 years). Children's psychosocial factors were parent-reported for children aged <12 years and self-reported by children aged ≥12 years.

*Adult participants*
*Depressive symptoms* were examined using the Beck Depression Inventory (BDI-II),(8) which is a self-report questionnaire consisting of 21 items that measure various symptoms of depression. We calculated a sum score (range 0-63 points) of depressive symptoms, where a cutoff of ≥14 points indicated the presence of depression.(8,9) The severity of depressive symptoms was classified as minimal (0-13 points), mild (14-19 points), moderate (20-28 points), or severe (29-63 points).(8) Adulthood *anxiety symptoms* were examined using the Generalized Anxiety Disorder-7 (GAD-7) scale.(10) We calculated a sum score (range 0-21 points), where a cutoff of ≥5 indicated presence of anxiety. The severity of anxiety symptoms was classified as minimal (0-4 points), mild (5-9 points), moderate (10-14 points), or severe (15-21 points).(10) Adulthood *quality of life* was examined using the World Health Organization's Quality of Life short measure (WHOQOL-BREF).(11) We used average scores from the psychological and social domains for adult participants across all generations.

*Child and adolescent participants*
*Depressive symptoms* in adolescents were examined using the Child Depression Inventory (CDI)(12) that was given to youth aged 12-17 years. We calculated a sum score of CDI (range 0-52). The cutoff for depression was defined based on sex-specific percentiles so that the proportion of depression cases corresponded to that based on BDI-II cases in adult males and females. In our data, the cut-off was 11 points for males (corresponding to the 91.8$^{th}$ percentile) and13 for females (corresponding to the 83.6$^{th}$ percentile). Children aged ≤11 years were not given depression questionnaires, but we used the Sadness dimension from the Rothbart Temperament in Middle Childhood Questionnaire(13,14) (aged 7-11 years) or from the Rothbart Child Behaviour Questionnaire(15) (aged 3-6 years) as a proxy for childhood depressiveness. We classified children who scored ≥85$^{th}$ percentile as having high depressive symptoms. *Anxiety-symptoms* in children and adolescents were examined using the Fear and Shyness temperament dimensions as proxies for the tendency to experience dispositional anxiety. Children aged 12-17 years completed self-report questionnaires on the Early Adolescent Temperament Questionnaire.(15) Children aged ≤11 years had their parents report on their behalf on the Temperament in Middle Childhood Questionnaire (children aged 7-11

years or the Child Behaviour questionnaire for children aged 3-6 years).(13,14) We classified children who scored ≥85th percentile as having high levels of anxiety-symptoms. *Quality of life* in adolescents (aged 7-11 years) was examined using the dimensions Self-esteem, Friends, Family and School from the Questionnaire for Measuring Health-Related Quality of Life in Children and Adolescents (KINDL).(16) Children aged ≤11 years were not presented any of the quality of life measures.

Each of the psychosocial outcomes (depressive symptoms, anxiety, quality of life) was harmonised across age groups to allow pooled analyses. Within each age group (G0, G1, G2 adults, G2 adolescents, G2 children 7-11 years of age, G2 children 3-6 years of age), the variables describing the outcomes were standardised to mean 100 and standard deviation 15 prior to pooling the data.

**Epigenetic markers as mediators**

*Semen samples*

Human semen was collected by masturbation (sexual abstinence for at least 48 hours recommended) from males aged 18 and older. The sample was transferred to an 8-ml tube containing 150 µl penicillin/streptomycin (Sigma, P4333) and 15 µl gentamicin sulphate (Sigma, G1397) and shipped to the site of research using a postal service. Upon arrival, the semen sample was stored at -20ºC until purification. For the purification of spermatozoa, the samples were thawed at room temperature and then incubated at +37ºC for 5-25 min for liquefaction. After counting the number of sperm, the ejaculate was pipetted on the top of 2 ml of 50% Puresperm (Nidacon) solution in a 15-ml conical tube (max 3 ml of the sample per one purification), and centrifuged at 400xg for 15 min. Five-hundred µl of the uppermost layer containing the semen plasma was pipetted into a 1.5-ml Eppendorf tube and stored at -20°C. The other layers were discarded, leaving around 1 ml including the spermatozoa in the pellet. The sperm pellet was washed by adding an equal volume of 2X Somatic Cell Lysis Buffer (final concentration 0.01% SDS, 0.05% Triton-X 100), and subsequent centrifugation at 3000xg for 15 min. The sperm pellet was further washed with 3 ml of phosphate-buffered saline (PBS) and centrifuged at 2500xg for 15 min. The sperm pellet was resuspended in 1 ml of PBS, and the number of sperm was counted. Six µl of the sperm sample was spread on an objective slide, air-dried, fixed with 95% ethanol for 15 min and stained with SPERM STAIN (Microptic). The slides were used to assess the purity of sperm samples and to calculate the percentage of somatic cell contamination. The remaining sperm suspension was divided into aliquots containing $1x10^7$ spermatozoa each. Spermatozoa were pelleted at 12 000xg for 5 min, the supernatant was discarded and sperm pellet was snap-frozen in liquid nitrogen and stored at -80°C.

*Sperm small-RNA-sequencing* - Flow-chart of the sperm small-RNA-sequencing is illustrated in Supplementary Figure 1. For RNA extraction, $1x10^7$ spermatozoa were thawed on ice, resuspended in 30 µl of nuclease-free water (NFW), and supplemented with 25 µl of 20 mg/ml solution of Proteinase K (Roche) and 50 µl of tris(2-carboxyethyl)phosphine (TCEP, Macherey-Nagel or Sigma-Aldrich, final concentration ~50 mM). After incubating at room temperature for 5 min, RNA was extracted with 1 ml TRIzol LS (Invitrogen) according to the manufacturer's instructions. After precipitation with isopropanol in the presence of 2 µl of GlycoBlue (15 mg/ml, Invitrogen) overnight at -20ºC, the RNA pellet was resuspended in NFW. After DNase I digestion (Sigma-Aldrich) without heat inactivation, RNA was re-extracted with Trizol LS, as described above. The precipitated RNA pellet was dissolved in 10 µl of NFW, and the quality of RNA was analysed by Bioanalyzer (Agilent RNA 6000 Pico Kit). Libraries for small-RNA-sequencing were prepared using NEB Next® Multiplex Small

RNA kit (New England Biolabs), and the libraries were sequenced by NovaSeq 6000 system (Illumina).

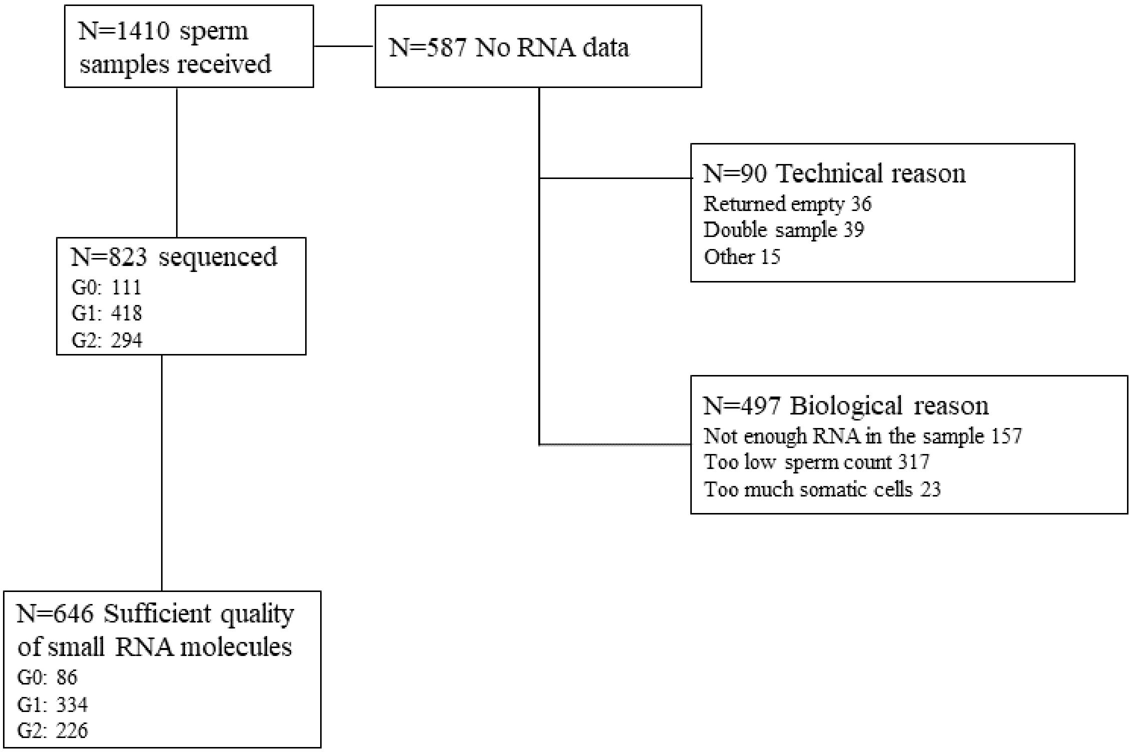


**Supplementary Figure 1.** Flow-chart of the sperm small-RNA-sequencing.

*Sperm DNA-methylation* - For sperm DNA methylation measurements, genomic DNA was isolated from pelleted sperm cells with modified protocol using Revvity Chemagic 360 robot with DNA-tissue 96 kit (art.no: Chemagic kit CMG-723). In short, the sperm cell pellets were incubated for one hour at RT in lysis buffer S (79.7µl) with 50 mM β- mercaptoethanol (0.34µl) and 20 mg/ml proteinase K (6µl). Then 120µl of lysis buffer B was added, 1h incubation repeated, and isolation performed with Chemagic 360 robot without adding the proteinase K and lysis buffer again. The quality of the samples was ensured using Agilent Bioanalyzer 2100 or Advanced Analytical Fragment Analyzer. Sample DNA concentration was measured with Qubit®/Quant-IT® Fluorometric Quantitation (Life Technologies), and/or Nanodrop ND-2000 (Thermo Scientific). Libraries for Reduced Representation Bisulfite Sequencing (RRBS) were prepared using Nugen Ovation RRBS Methyl-Seq with TrueMethyl BS (Tecan), and the libraries were sequenced by NovaSeq 6000 system (Illumina).

### Other variables and covariates

*Blood sampling and biochemical measurements*

Venous blood samples were taken after a minimum of a 4-hour fast and stored at -70°C until analysis. Creatinine, and liver enzymes were determined from serum samples with enzymatic assays using the following system reagents; Creatinine (enzymatic), ALT/GPT (IFCC, with pyridoxal-5-phosphate activation), AST/GOT (IFCC, with pyridoxal-5-phosphate activation), and Gamma-GT (IFCC) (all from Thermo Fisher Scientific, Finland) on an Indiko Plus analyzer (Thermo Fisher Scientific, Finland). Apolipoprotein A1, Apolipoprotein B, Lipoprotein(a), and C-reactive protein (CRP) were determined from serum samples and glycated hemoglobin (HbA1c) from hemolyzed whole-blood with turbidimetric assays on the Indiko Plus analyzer using system reagents; Apolipoprotein A1, Apolipoprotein B, Lipoprotein(a), CRP High Sensitivity and HbA1c (all from Thermo Fisher Scientific, Finland). The range for detection of CRP concentration was 0.25-10 mg/l. Concentrations exceeding the upper limit of the range were determined as 10.5 mg/l and the concentrations below the lower limit were determined as 0.125 mg/l.

*Serum metabolomics* - A high-throughput nuclear magnetic resonance (NMR) spectroscopy metabolomics platform (17) was used to quantify 164 lipid and metabolite measures from serum.(18) The platform applies a single experimental setup, which allows for simultaneous quantification of standard clinical lipids, 14 lipoprotein subclasses and individual lipids (triglycerides, phospholipids, free and esterified cholesterol) transported by these particles, multiple fatty acids, glucose and various glycolysis precursors, ketone bodies and amino acids in absolute concentration units.

*Blood DNA methylation* - Blood DNA methylation profiling was performed from leukocytes obtained from 1-4 ml of EDTA blood samples using a Perkin Elmer Chemagic (CMG-1074) according to the manufacturer's instructions, with the exemption of utilizing modified binding buffer 2 with a subset of the samples. Profiling was performed with the Illumina Infinium MethylationEPIC BeadChip at Helmholtz Zentrum, Munich, Germany. Samples were applied to the arrays in a randomised order. Aliquots of 1 µg of DNA were subjected to bisulphite conversion, and a 4-µl aliquot of bisulphite-converted DNA was subjected to whole-genome amplification, followed by enzymatic fragmentation and hybridization onto an Illumina Infinium MethylationEPIC BeadChip (version 1 and 2). The arrays were scanned with the iScan reader (Illumina). Background subtraction was performed on all probe values using the bgcorrect.illumina function in the minfi package. Probes with a detection p-value of higher than $1x10^{-16}$ and samples with a sample call rate of more than 95% were filtered out. Autosomal probes were separated into six groups by probe-type, and quantile normalization was performed separately for each group. Samples for which the actual sex did not match the predicted sex were excluded and quality control was performed on the data. All pre-processing steps were performed using functions implemented in the minfi, sesame or limma R/Bioconductor package.

*Genome-wide genotyping* **-** Genome-wide genotyping was done using Illumina Human670-Quad Custom (G1) and Illumina Infinium Global Screening Array (G0, G2 and a subset of G1). Genotypes were called using Illumina's GenCall algorithm. The following filters were used for sample and SNP quality control (QC): sample and SNP call rate <0.95, cryptic relatedness (pi-hat >0.2), SNP Hardy-Weinberg equilibrium test ($p \leq 1x10^{-6}$). Samples with sex discrepancy, excess heterozygosity, as well as genetic outliers detected with multidimensional scaling (MDS), were excluded. Genotype imputation was performed using Minimac3 and TOPMed r3 reference set on the TOPMed Imputation Server.

*MicroRNA profiling* - MicroRNA expression profiling was performed from serum samples. MicroRNAs were isolated from 200 µl of serum with the miRNeasy Serum/Plasma Kit (Qiagen), including the DNase Set, using the QiaCube. Profiling was performed with a custom TaqMan® OpenArray® MicroRNA Panel (Life Technologies), with 168 microRNAs shown to be widely expressed in serum. The scaled-down Low Sample Protocol was used. Only assays with an Amplification score of >1 and Cq Confidence of >0.7 were accepted. The data were normalised with a global mean normalization approach in R language. The approach involves the calculation of a normalization factor as the global mean of all expressed microRNAs per sample. Furthermore, between-sample normalization was carried out with the quantile normalization method. Batch correction, when needed, was performed with ComBat.(19)

*Faecal samples and metagenome sequencing*
The faecal samples were collected using commercial, validated, state-of-the-art microbial collection kits (OMNIgene-Gut 200, DNA Genotek) by the participants at home. Parents were advised to assist their children in sample collection if needed. A similar methodology has been applied by well-established groups in the research field.(20) The samples were returned via mail to the laboratory where they were homogenised, divided into three aliquots and stored at -80°C. The participants reported on the use of antimicrobials, probiotics, and stool consistency at the time of sampling. DNA from the faecal samples was extracted in 96-well format using DNA Stool 200 Kit special H96 (PerkinElmer, Finland) kit with a Magnetic Separation Module I automated instrument in the Turku Microbiome Biobank. Prior to the extraction, a mechanical lysis step by bead-beating 15 Hz for 5 min x2 (TissueLyser II, Qiagen) was performed. The obtained DNA concentration was measured with a Qubit 2.0 Fluorometer (Thermo Fisher Scientific, USA) using a Qubit dsDNA High Sensitivity Assay kit. DNA integrity was determined by 1% TBE agarose gel. The DNA was divided into two 100-µL aliquots and stored at -80°C. Deep shotgun metagenomics sequencing will be performed using NovaSeq 6000 system (Illumina) to obtain DNA sequence data ≥6 Gb/sample to reach strain-level taxonomy.(21)

*Urine samples and metabolomics*
Urinary samples were collected in one of the participating centres (Tampere) covering all three generations. The sample was first collected to a urine cup. Five ml of urine was immediately transferred to a 5 ml tube and stored in -80 C. Urine samples were analysed from those participants who had also given serum sample and provided clinical data.(22) A total of 43 urinary metabolites were quantified with an open-access methodology for high-throughput urinary NMR metabolomics.(23)

*Longitudinal movement of the common carotid arterial wall*
To assess the longitudinal movement of the common carotid arterial wall, participants were advised to stop breathing for 5 seconds and a simultaneous clip was recorded and stored for subsequent off-line analysis. Carotid artery longitudinal wall motion analysis is performed using an in-house motion tracking program (24) in line with the practical guidelines.(25) The software written in MATLAB (The MathWorks Inc., Natic, MA, USA), is capable of reading the graphical electrocardiogram information of the ultrasound recording and simultaneously tracking the longitudinal and radial motions of the arterial wall. The basic method used in the motion tracking is a two-dimensional cross-correlation (block matching) enhanced with a contrast optimization technique to reduce noise from video.(25)

*Socioeconomic status*
Data on education, income and occupational status were collected from participant self-report. Level of basic education was queried using six categories. Educational attainment after basic education was queried using 11 categories and as total years of education. Participant's own gross annual income was measured on a 21-point scale ranging from 0 (<5 000€) to 21 (>100 000€). In addition, gross yearly income of the participant's household was measured on a 26-point scale ranging from 0 (<5 000€) to 26 (>150 000€). Occupational status was self-reported using six categories. Moreover, participants were asked to report their current occupation, and if not working then the latest occupation. Finally, the position of leadership was queried using four categories ranging from 'no' to 'yes, and my subordinate's subordinate has subordinates.

*Lifestyle factors*
Data on food consumption, smoking habits, alcohol consumption and physical activity of the participants were collected by self-report questionnaires.

*Food consumption* - Food consumption during the past 12 months was assessed by a validated quantitative 134-item food frequency questionnaire (FFQ) from all generations. The questionnaire was slightly modified for the participants aged < 12 years by omitting the questions concerning alcohol use. The average use of the food items and mixed dishes was recorded by ten frequency categories ranging from never or seldom to at least six times a day. The average daily intake of food groups (e.g., vegetables, rye and oil) and nutrients (e.g., fibre and vitamin C) were estimated by multiplying the fixed portion sizes with food consumption frequency. The portion size was fixed for each food item or mixed dish (e.g., slice and glass) based on the dietary interviews of the national FINDIET 2007 Study.(26,27) The National Food Composition Database (Fineli®) maintained by the Finnish Institute for Health and Welfare was utilised when calculating energy and nutrient intakes. Exclusions were made due to incompletely filled FFQs and daily energy intake cut-off points corresponding to 0.5% at both ends of the daily energy intake distributions for males and females.

*Smoking habits* - Smoking habits were queried from all participants aged 12 years and older who reported that they had ever smoked. The participants were asked to report at which age they began smoking and if ceased, at what age they ceased. Current smoking was queried with six categories. Participants were asked to report the number of years they have smoked daily, and the number of tobacco products consumed per day. Additionally for e-cigarettes, amount of weekly use of liquid and nicotine concentration of the liquid were queried. Passive smoking exposure was queried using 5 categories ranging from 0h to ≥4h.

*Alcohol consumption* - Alcohol consumption was queried from all participants aged 12 years and older who reported that they had ever drank at least one portion of alcohol. Frequency of alcohol drinking was queried using eight categories ranging from never to daily or almost daily. Number of portions consumed per drinking session was assessed as self-reported consumption of 1/3 litre cans or bottles of beer, glasses (12 cl) of wine, and 4 cl shots of liquor or strong alcohol (these portions are comparable to c. 14 g of alcohol). Participants were asked to report the number of portions of above-mentioned alcohol beverages usually consumed per week. Frequency of binge drinking (≥6 portions per drinking session) was queried using 6 categories ranging from 0-1 times per year to at least twice per week. Additionally, number of times the participant had hangover during past 12 months was queried using six categories ranging from 0-1 times to once a week or more.

*Physical activity* - Data on physical activity was queried with questionnaires including items on the frequency and intensity of leisure-time physical activity/sports, participation in organised sports/physical activity, active commuting to school/work, occupational PA and sedentary time. In addition to questionnaires, physical activity was measured with a validated motion sensor, i.e., triaxial ActiGraph accelerometer (GT3X+ and wGT3X+; ActiGraph, Pensacola, FL). The device was attached to a waistband or belt, and the motion data were collected for seven consecutive days. Participants were instructed to wear accelerometers also when sleeping, but to take device off for bathing and water activities. Participants kept a diary in which they entered their sleeping and working periods, an estimation of sleeping time, and whether anything extraordinary, such as illness, occurred during the measurement period. Data were collected at a 60-Hz sample rate using normal filter and later averaged to 60-s epochs. A total of 60 minutes of consecutive zero counts were defined as non-wearing time and excluded from data. Individual-level data from at least ≥10 hours for ≥4 days during awake time were considered valid. The variables to be calculated from accelerometer data include total physical activity, steps/day, time spent sedentary, light, moderate and vigorous intensity physical activity.

*Additional psychosocial measurements*
*Adulthood - Personality characteristics* were assessed using the Neuroticism-Extraversion-Openness-Five-Factor-Inventory,(28,29) the Minnesota Multiphasic hostility questionnaire,(30) the Emotionality-Activity-Sociability questionnaire,(31) the Rational-Experiential cognitive style Inventory,(32) and the Life-Orientation Test questionnaire.(33) *Social relations* were assessed using the Multidimensional Scale of Perceived Social Support scale,(34) the Sarason Social Support scale,(35) and the short version of the UCLA Loneliness scale.(36) *Sleep patterns* were assessed using the Epworth Sleepiness scale(37) and the Circadian preference scale.(38) *Psychosocial work environment* was assessed using the Job Content Questionnaire(39) and the short version of the Colquitt Organizational Justice questionnaire.(40) Student participants filled in a three-item student burnout questionnaire.(41) *Sleep quality* was assessed using the Jenkins Sleep problems scale,(42) which was dichotomised based on age-specific 75$^{th}$ percentile.

*Childhood and adolescence - Temperament characteristics* were examined using the Emotionality-Activity-Sociability questionnaire,(43) the Children's Behavior Questionnaire Short Form(44) (<7 years of age), Temperament in Middle Childhood Questionnaire(45) (7-11 years of age) and Early Adolescent Temperament Questionnaire(15) (≥12 years of age). *Social relations* of the adolescent participants (≥12 years) were assessed using the Multidimensional Scale of Perceived Social Support scale,(34) the Sarason Social Support scale,(35) and the short version of the UCLA Loneliness scale,(36) while social relations of the participants <12 years were assessed using the HOME Short Form scale.(46) *Sleep quality* was assessed using the Jenkins Sleep problems scale(42) for children and adolescents of all ages. School-related problems were assessed by asking specific questions of school burnout and school achievements from the adolescent participants.

*Pubertal status*
Self-rated pubertal status was determined from all participants aged 8-17 years. The participants were shown a sex-specific illustration presenting pubertal stages according to the Tanner staging. Both sexes reported their pubic hair development (P; stages 1-6, 1 indicating no puberty and 6 indicating completed puberty). Female participants reported additionally breast development (B; stages 1-5, 1 indicating no puberty and 5 indicating completed puberty), while male participants reported development of external genitalia (G; stages 1-5, 1 indicating no puberty and 5 indicating completed puberty). For females, onset of puberty was considered to occur when breast development was reported to be at stage breast 2 (B2) and pubic hair at stage 2 (P2). For males, external genitalia at stage 2 (G2) and public hair at stage 2 (P2) was considered to indicate the onset of puberty.

*Child health clinic cards*
In Finland all children under school age are entitled to child health clinic services since birth. The data on child's health, growth and development are recorded in a health clinic card. G1 participants were asked to bring their child health clinic card to the study visit if possible. The cards were scanned and information on height and weight development was recorded.

**References**


1. Hernesniemi JA, Heiskanen J, Ruohonen S, Kartiosuo N, Hutri-Kähönen N, Kähönen M, et al. Aortic sinus diameter in middle age is associated with body size in young adulthood. *Heart* 2018;104:773-778.

2. Heiskanen JS, Hernesniemi JA, Ruohonen S, Hutri-Kähönen N, Kähönen M, Jokinen E, et al. Influence of early-life body mass index and systolic blood pressure on left ventricle in adulthood - the Cardiovascular Risk in Young Finns Study. *Ann Med* 2021;53:160-168.

3. Cole TJ, Bellizzi MC, Flegal KM, Dietz WH. Establishing a standard definition for child overweight and obesity worldwide: International survey. *BMJ (Clinical Research Ed.)* 2000;320:1240-1243.

4. Friedewald WT, Levy RI, Fredrickson DS. Estimation of the concentration of low-density lipoprotein cholesterol in plasma, without use of the preparative ultracentrifuge. *Clin Chem* 1972;18:499-502.

5. Raitakari OT, Juonala M, Kähönen M, Taittonen L, Laitinen T, Mäki-Torkko N et al. Cardiovascular risk factors in childhood and carotid artery intima-media thickness in adulthood: the Cardiovascular Risk in Young Finns Study. *JAMA* 2003;290:2277-2283.

6. Touboul P-J, Hennerici MG, Meairs S, Adams H, Amarenco P, Bornstein N et al. Mannheim Carotid Intima-Media Thickness and Plaque Consensus (2004–2006–2011). An update on behalf of the advisory board of the 3rd, 4th and 5th Watching the Risk Symposia, at the 13th, 15th and 20th European Stroke Conferences, Mannheim, Germany, 2004, Brussels, Belgium, 2006, and Hamburg, Germany, 2011. Cerebrovasc Dis 2012;34:290–296.

7. Flury BN. Common principal components in k groups. J Am Stat Assoc 1984;79:892-898.

8. Beck AT, Steer RA, Brown GK. BDI-II Beck Depression Inventory. San Antonio: The Psychological Corporation; 1996.

9. Viinamäki H, Tanskanen A, Honkalampi K, Koivumaa-Honkanen H, Haatainen K, Kaustio O et al. Is the Beck Depression Inventory suitable for screening major depression in different phases of the disease? *Nordic J Psychiatry* 2004;58:49-53.

10. Spitzer RL, Kroenke K, Williams JB, Löwe B. A brief measure for assessing generalized anxiety disorder: the GAD-7. *Arch Intern Med* 2006;166:1092-1097.

11. Development of the World Health Organization WHOQOL-BREF quality of life assessment. The WHOQOL Group. *Psychol Med* 1998;28:551-558.

12. Kovacs M. The Children's Depression Inventory (CDI). *Psychopharmacol Bul* 1985;21:995-998.

13. Nyström B, Bengtsson H. A psychometric evaluation of the Temperament in Middle Childhood Questionnaire (TMCQ) in a Swedish sample. *Scand J Psychol* 2017;58:477-484.

14. Simonds J. The role of reward sensitivity and response: Execution in childhood extraversion. University of Oregon 2006.

15. Capaldi DM, Rothbart MK. Development and Validation of an Early Adolescent Temperament Measure. *J Early Adolesc* 1992;12:153-173.

16. Ravens-Sieberer U, Bullinger M. Assessing health-related quality of life in chronically ill children with the German KINDL: first psychometric and content analytical results. *Qual Life Res* 1998;7:399-407.

17. Mäkinen VP, Kettunen J, Lehtimäki T, Kähönen M, Viikari J, Perola M et al. Longitudinal metabolomics of increasing body-mass index and waist-hip ratio reveals two dynamic patterns of obesity pandemic. *Int J Obes* 2023;47:453-462.

18. Soininen P, Kangas AJ, Würtz P, Tukiainen T, Tynkkynen T, Laatikainen R et al. High-throughput serum NMR metabonomics for cost-effective holistic studies on systemic metabolism. *Analyst* 2009;134:1782-1785.

19. Johnson WE, Li C, Rabinovic A. Adjusting batch effects in microarray expression data using empirical bayes methods. *Biostatistics* 2007;8:118–127.

20. de Goffau MC, Jallow AT, Sanyang C, Prentice AM, Meagher N, Price DJ, et al. Gut microbiomes from Gambian infants reveal the development of a non-industrialized Prevotella-based trophic network. *Nat Microbiol* 2022;7:132–144.

21. Isokääntä H, Tomnikov N, Vanhatalo S, Munukka E, Huovinen P, Hakanen AJ, et al. High-throughput DNA extraction strategy for fecal microbiome studies. *Microbiol Spectr* 2024;12:e0293223.

22. Li T, Ihanus A, Ohukainen P, Järvelin M-R, Kähönen M, Kettunen J et al. Clinical and biochemical associations of urinary metabolites: quantitative epidemiological approach on renal-cardiometabolic biomarkers. *Int J Epidemiol* 2024;53:dyad162.

23. Tynkkynen T, Wang Q, Ekholm J, Anufrieva O, Ohukainen P, Vepsäläinen J et al. Proof of concept for quantitative urine NMR metabolomics pipeline for large-scale epidemiology and genetics. *Int J Epidemiol* 2019;48:978–993.

24. Yli-Ollila H, Laitinen T, Weckström M, Laitinen TM. Axial and radial waveforms in common carotid artery: An advanced method for studying arterial elastic properties in ultrasound imaging. *Ultrasound Med Biol* 2013;39:1168–1177.

25. Rizi FY, Au J, Yli-Ollila H, Golemati S, Makūnaitė M, Orkisz M, et al. Carotid wall longitudinal motion in ultrasound imaging: An expert consensus review. *Ultrasound Med Biol* 2020;46:2605–2624.

26. Pietinen P, Paturi M, Reinivuo H, Tapanainen H, Valsta LM. FINDIET 2007 Survey: energy and nutrient intakes. *Public Health Nutr* 2010;13:920-924.

27. Reinivuo H, Hirvonen T, Ovaskainen ML, Korhonen T, Valsta LM. Dietary survey methodology of FINDIET 2007 with a risk assessment perspective. *Public Health Nutr* 2010;13:915-919.

28. McCrae RR, Costa PT. A contemplated revision of the NEO Five-Factor Inventory. *Pers Indiv Diff* 2004;36:587-596.

29. Costa PT Jr, McCrae RR. Revised NEO Personality Inventory (NEO-PI-R) and NEO Five Factor Inventory (NEO-FFI) professional manual. Odessa, FL: Psychological Assessment Resources; 1992.

30. Hathaway SR, Mc Kinley JC. A multiphasic personality schedule (Minnesota): I. Construction of the schedule. *J Psychol* 1940;10:249-254.

31. Buss AH. The EAS theory of temperament. In: Strelau J, Anglneitner A, eds. Explorations in temperament. New York: Plenum Press; 1991:43-60.

32. Epstein S, Pacini R, Denes-Raj V, Heier H. Individual differences in Intuitive-Experiential and Analytical-Rational Thinking Styles. *J Pers Soc Psychol* 1996;71:390-405.

33. Scheier MF, Carver CS, Bridges MW. Distinguishing optimism from neuroticism (and trait anxiety, self-mastery, and self-esteem): A reevaluation of the Life Orientation Test. *J Pers Soc Psychol* 1994;67:1063-1078.

34. Zimet GD, Dahlem NW, Zimet SG, Farley GK. The Multidimensional Scale of Perceived Social Support. *J Pers Assessment* 1988;52:30-41.

35. Sarason I, Levine H, Basham R, Sarason B. Assessing social support: the social support questionnaire. *J Pers Soc Psychol* 1983;44:127-139.

36. Hughes ME, Waite LJ, Hawkley LC, Cacioppo JT. A short scale for measuring loneliness in large surveys: Results from two population-based studies. *Res Aging* 2004;26:655-672.

37. Johns MW. A new method for measuring daytime sleepiness: the Epworth sleepiness scale. *Sleep* 1991;14:540-545.

38. Horne JA, Ostberg O. A self-assessment questionnaire to determine morningness-eveningness in human circadian rhythms. *Int J Chronobiol 1975;*4:97e110.

39. Karasek R, Brisson C, Kawakami N, Houtman I, Bongers P, Amick B. The Job Content Questionnaire (JCQ): an instrument for internationally comparative assessments of psychosocial job characteristics. *J Occup Health Psychol* 1998;3:322-355.

40. Elovainio M, Heponiemi T, Kuusio H, Sinervo T, Hintsa T, Aalto AM. Developing a short measure of organizational justice: A multisample Health Professionals Study,. *J Occup Environ Med* 2010;52:1068-1074.

41. Salmela-Aro K, Kiuru N, Leskinen E, Nurmi J-E. School Burnout Inventory: Reliability and validity. *Eur J Psychol Assesment* 2009;25:48-57.

42. Jenkins DC, Stanton B-A, Niemcryk SJ, Rose RM. A scale for the estimation of sleep problems in clinical research. *J Clin Epidemiol* 1988;41:313-321.

43. Buss AH, Plomin R. Temperament: Early developing personality traits. Hillsdale (NJ): Erlbaum; 1984.

44. Rothbart MK, Ahadi SA, Hershey KL, Fisher P. Investigations of temperament at three to seven years: the Children's Behavior Questionnaire. *Child Development* 2001;72:1394-1408.

45. Ellis LK, Rothbart M. Early Adolescent Temperament Questionnaire Revised Short Form. Translated by Katri Räikkönen-Talvitie, University of Helsinki 1999.

46. Caldwell BM, Bradley RH. Home Inventory Administration Manual, Third Edition. In: Little Rock, AR. University of Arkansas at Little Rock; 2001.

**Supplementary Table 2**. Age-group specific numbers (% of all invited / % of those who attended the study visit) of provided data among the offspring of the original YFS participants (G2).

| | ≥18 years | 13-17 years | 7-12 years | 3-6 years |
|---|---|---|---|---|
| | **N (%)** | **N (%)** | **N (%)** | **N (%)** |
| **Questionnaires**[a] | | | | |
| **Basic** | 1504 (48.7)<br>1444 (97.8) | 658 (42.9)<br>576 (93.2) | 398 (47.3)<br>347 (95.3) | 92 (39.5)<br>76 (89.4) |
| **Diet** | 1310 (42.4)<br>1265 (85.7) | 490 (31.9)<br>464 (75.1) | 297 (35.3)<br>283 (77.8) | 69 (29.6)<br>66 (77.7) |
| **Psychosocial wellbeing** | 1327 (43.0)<br>1283 (86.9) | 504 (32.9)<br>478 (77.4) | 300 (35.7)<br>285 (78.3) | 71 (30.5)<br>68 (80.0) |
| **Samples** | | | | |
| **Semen**[b] | 388<br>(24.3 / 63.2) | na. | na. | na. |
| **Blood** | 1459<br>(47.3 / 98.9) | 565<br>(36.8 / 91.4) | 299<br>(35.6 / 82.1) | 67<br>(28.8 / 78.8) |
| **Faecal** | 803<br>(26.0 / 54.4) | 217<br>(14.2 / 35.1) | 172<br>(20.5 / 47.3) | 54<br>(23.2 / 63.5) |
| **Hair** | 1045<br>(33.8 / 70.8) | 428<br>(27.9 / 69.3) | 277<br>(30.9 / 76.1) | 65<br>(28.0 / 76.5) |
| **Anthropometrics**[c] | 1474<br>(47.7 / 99.9) | 617<br>(40.2 / 99.8) | 362<br>(43.0 / 99.5) | 84<br>(36.1 / 98.8) |
| **Blood pressure** | 1470<br>(47.6 / 99.6) | 613<br>(40.0 / 99.2) | 360<br>(42.8 / 98.9) | 81<br>(34.8 / 95.3) |
| **Ultrasonography** | | | | |
| **Vascular**[d] | 1473<br>(47.7 / 99.8) | 615<br>(40.1 / 99.5) | 355<br>(42.2 / 97.5) | 81<br>(34.8 / 95.3) |
| **Liver**[e] | 1473<br>(47.7 / 99.8) | 615<br>(40.1 / 99.5) | 356<br>(42.3 / 97.8) | 81<br>(34.8 / 95.3) |
| **Cognitive function** | 1466<br>(47.5 / 99.3) | 608<br>(39.6 / 98.4) | 359<br>(42.7 / 98.6) | 85<br>(36.5 / 100) |

[a]for questionnaires, the first numbers and proportions (%) are of all invited, and the second numbers and proportions (%) are of those who attended the study visit
[b]samples including sperms, proportion of men who were aged ≥18 years at the beginning of study/reached the age by the study visit
[c]numbers/proportions of those with height and/or weight
[d]numbers/proportions of those with any vascular imaging data
[e]numbers/proportions of those with successfully obtained fatty liver data
