## Supplementary material for "Cohort Profile Update: Expanding the Cardiovascular Risk in Young Finns Study into a multigenerational cohort": Statistical analysis plan

### 1. INTRODUCTION

This statistical analysis plan is a supplement to the article *Cohort Profile Update: Expanding the Cardiovascular Risk in Young Finns Study into a multigenerational cohort*. The plan has been created to investigate the original hypotheses listed in the research plan of the MULTIEPIGEN project (https://cordis.europa.eu/project/id/742927). The MULTIEPIGEN examines whether paternal exposures to certain stressors are linked with offspring phenotypes mediated by epigenetic markers in the sperm.

Experimental animal studies have provided powerful evidence indicating that certain epigenetic markers in sex cells can transfer information from fathers to the offspring. In humans, however, it is not possible to perform multigenerational controlled trials to test causal epigenetic relations between parental environmental exposures and offspring. Therefore, the only way to address this question is by using observational epidemiologic study designs. The MULTIEPIGEN is the first *a priori* designed epidemiologic study assessing the role of early life exposures, including chemical and psychosocial stress, in the development of cardio-metabolic, cognitive and psychosocial outcomes in their offspring.

The MULTIEPIGEN examines pre-specified ancestral exposures that based on animal studies have very high plausibility causing intergenerational effects on offspring phenotypes, including obesity-related phenotypes (components of the metabolic syndrome), cognitive function and psychological well-being. These exposures include ancestral stressors, such as tobacco smoke, obesity, organic pollutants, and accumulation of psychosocial adversities.

There are no standard approaches for analysing such multigenerational mediation questions. The purpose of this document is to pre-specify the key principles of multigenerational analyses that will be implemented in the MULTIEPIGEN project.

#### 1.1 Conceptual framework

Figure 1 presents the conceptual framework of multigenerational inheritance over three generations as a directed acyclic graph (DAG). Each arrow indicates a hypothesised causal association. Two defined sets of paths constitute the two key hypotheses on epigenetic inheritance in the MULTIEPIGEN project: intergenerational and transgenerational epigenetic inheritance (Figures 2 and 3). In practice, our plan is to investigate the two pathways of epigenetic inheritance using a mediation analysis approach, described in more detail below (see Section 4.1, *'Mediation analysis'*).

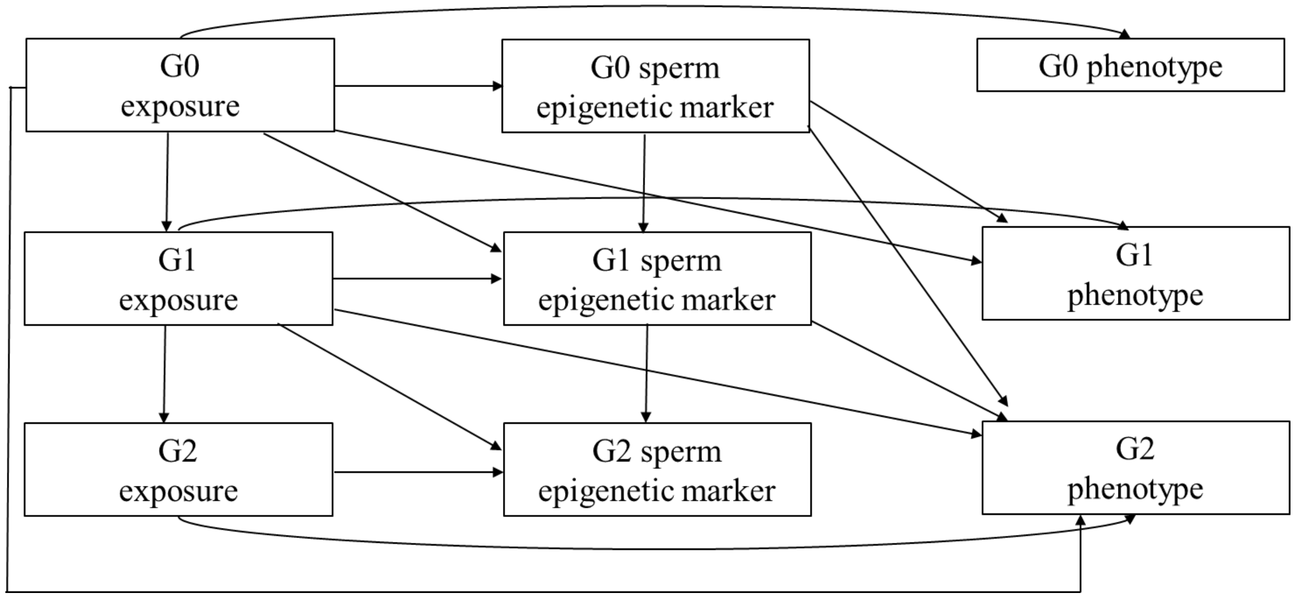


*Figure 1. The multigenerational framework in the MULTIEPIGEN project. Each arrow represents a hypothesised causal or statistical association. Within individual, exposure levels could affect the phenotypes and sperm epigenome (horizontal arrows). Some exposures, such as cigarette smoking, correlate over the generations (left-hand side vertical arrows). Sperm epigenetic markers might be correlated over the generations in the male line (middle vertical arrows). The diagonal arrows represent inter- and transgenerational effects. The intergenerational and transgenerational hypotheses are detailed in Figures 2 and 3.*

*Intergenerational epigenetic inheritance*

By intergenerational epigenetic inheritance, we refer to a transmission of the effect of paternal exposure to an environmental stressor to the subsequent generation via the paternal sperm epigenome. Figure 2 highlights the pathways corresponding to such intergenerational epigenetic inheritance, separately for the G0–G1 and G1–G2 dyads. In this hypothesis, the father (i.e., G0 in the G0–G1 dyad and G1 in the G1–G2 dyad) and his sperm cells are exposed to the environmental stressor prior to the conception of the offspring (i.e., G1 in the G0–G1 dyad and G2 in the G1–G2 dyad). The subsequent generation is considered to be intergenerationally exposed via the paternal sperm cells impacted by the environmental stressor.

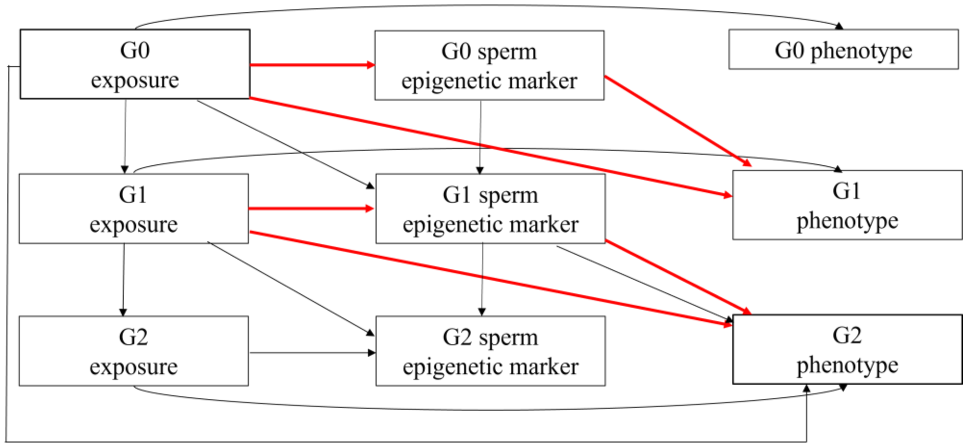


*Figure 2. The hypothesis on intergenerational epigenetic inheritance. The red arrows describe the two sets of pathways for intergenerational epigenetic inheritance investigated in the three-generational data: one for G0—G1 dyads and one for G1—G2 dyads. The outcomes, i.e., G1 and G2 phenotypes, are measured from both male and female participants.*

*Transgenerational epigenetic inheritance*
The transgenerational epigenetic inheritance hypothesis builds upon the intergenerational hypothesis and extends it to three successive generations. The framework of transgenerational epigenetic inheritance over G0–G1–G2 generations is emphasised in Figure 3. In this framework, the G0 grandfathers and their sperm cells are exposed to the stressor, while the reproductive cells of the G1 are unexposed. For example, in the context of smoking this means that the analyses are confined to non-smoking G1 fathers. The health consequences of the environmental stressor persist to the G2 phenotypes, i.e., the G0 exposure affects the G2 phenotype.

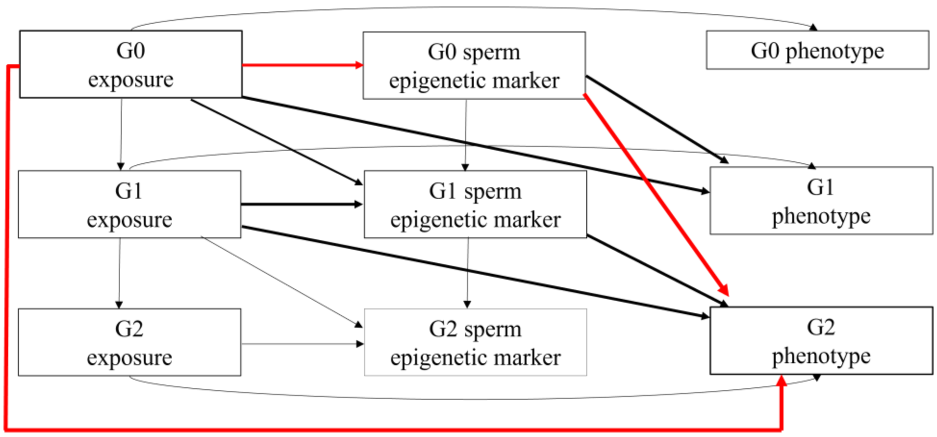


*Figure 3. The hypothesis on transgenerational epigenetic inheritance. The red arrows describe the pathways from G0 to G2 via transgenerational epigenetic inheritance. The bolded black arrows refer to intergenerational epigenetic inheritance. The outcomes, i.e., G2 phenotypes, are measured from both male and female participants. In addition to male G1 participants, a similar scheme can be extended to maternal grandfathers.*

### 2. EXPOSURES, OUTCOMES, MEDIATORS, AND COVARIATES

*Exposures*
The pre-specified paternal and grandpaternal exposures of interest are assessed for generations G0 and G1. The primary exposures are:
- Smoking at different stages during life-course and the course of development and childhood of the offspring
- Psychosocial adversities at different life-course stages and stages during the development of the offspring
- Life-course adiposity
- Serum levels of persistent organic pollutants (only available for G1, i.e., for the G1–G2 intergenerational hypotheses) measured at three time points (the baseline (1980); youth (1992); and early adulthood (2001)). The pollutants include organochlorine compounds, polychlorinated biphenyls, polybrominated biphenyls and perfluoroalkyl and polyfluoroalkyl substances.

*Outcomes*
The pre-specified offspring outcomes of interest are:
- Cardio-metabolic phenotypes: BMI, waist circumference, blood pressure (systolic and diastolic), total cholesterol, HDL-cholesterol, LDL-cholesterol, apolipoprotein B, apolipoprotein A-1, triglycerides (ln-transformed), glucose, insulin (ln-transformed), hepatic steatosis, carotid artery intima-media thickness and elasticity, and carotid artery plaques

- Cognitive function domains: episodic memory and associative learning, working memory, executive function, reaction and movement time, sustained attention
- Psychological wellbeing: quality of life, depression, anxiety, and quality of sleep.

The outcomes are assessed from all generations.

*Mediators*

The mediators of interest are assessed from G0 and G1 sperm samples: (a) sperm non-coding RNA profile, consisting of microRNAs, transfer RNAs-derived fragments, and piwi-interacting RNA clusters and sequences; (b) sperm DNA methylation markers. Potential mediators are be identified from a large set of RNA molecules and DNA methylation markers as described in Section 4 (Statistical Methods).

*Adjustment for confounding*

Mediation analysis is subject to assumptions of no unmeasured confounding of the mediator-outcome, exposure-outcome and exposure-mediator associations and controlling for potential confounding variables is thus of high importance. Each analysis will be controlled for potential confounders, including but not necessarily limited to the socio-economic status, paternal age, offspring age, and offspring sex.

In the original research plan, only exposure to tobacco smoke, persistent organic pollutants and psychosocial adversities were listed as paternal exposures, and only the sperm non-coding RNA molecules as potential epigenetic mediators. Due to recent advancements in the field, the research plan for the original hypotheses has required amendments, as a result, we have added paternal adiposity as a hypothesised exposure, and semen DNA methylation as a hypothesised mediator (1–3).

In addition to the predefined variables and their roles listed above, we acknowledge that the field of epigenetic inheritance is advancing and thus considering additional variables might be necessary. Therefore, to complement the analyses described in this statistical analysis plan, guided by the predefined alternatives of ancestral exposure, epigenetic mediators, and offspring outcomes, we will consider performing other analyses using different available exposure, mediator and outcome variables in case such analyses can be justified by future scientific advancements in this field or otherwise considered necessary.

### 3. ANALYSIS DATA SETS

The sample sizes are reported in the main text of the cohort profile. To maximally utilise the collected data, each individual association/pathway of the DAGs will primarily be analysed using the full analysis set, i.e., utilising information from all participants who have the required information. This means, for example, that all participants with data on exposure and the sperm epigenetic profile will be included in the epigenome-wide association study (EWAS) analyses, regardless of whether they have (data on) offspring. However, the mediation analyses will be implemented on complete cases only, i.e., on those dyads with data on (grand)paternal (G0 or G1) exposure and sperm epigenome as well as on offspring (G1 or G2) phenotype.

### 4. STATISTICAL METHODS

#### 4.1 Mediation analysis

Mediation analysis is a family of statistical methods to quantify the causal effect of a specified exposure on a specified outcome and the extent to which this effect is transmitted via an intermediate variable, a mediator (4). Here, we first present the general idea of mediation analysis and then describe how the hypotheses of intergenerational and transgenerational epigenetic inheritance will be addressed as mediation problems. Figure 4 presents a DAG describing mediation with a single mediator. The exposure affects the outcome via two pathways: directly, i.e., independently of the mediator, and indirectly, i.e., via its effect on the mediator and the subsequent effect of the mediator on the outcome.

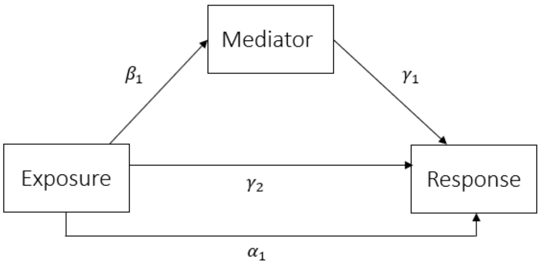


*Figure 4. A DAG describing a general mediation question with a single mediator.*

In practice, mediation analysis is based on two regression models describing the dependencies between the exposure, mediator and outcome:

$$E[Y|x,m,c] = \gamma_0 + \gamma_1 m + \gamma_2 x + \gamma_3' c \quad (1)$$

$$E[M|x,c] = \beta_0 + \beta_1 x + \beta_2' c, \quad (2)$$

where $X$ denotes the exposure, $M$ the mediator, $Y$ the outcome, and C the set of covariates adjusted for (not shown in the DAG of Figure 1) (4). In our context, $X$ corresponds to one of the paternal or grandpaternal exposure variables, M to the paternal or grandpaternal sperm epigenetic marker, and Y to one of the offspring phenotypes. The model for $Y$ can be extended to cover logistic regression for binary phenotypes, such as dichotomised cardio-metabolic phenotypes, existence of plaques in carotid arteries, or depression. (4)

In the above model, parameter $\gamma_2$ is the direct effect of the exposure on the outcome, i.e., the effect not due to mediation. The parameters of $\beta_1$ (the effect of exposure on mediator) and $\gamma_1$ (the effect of mediator on outcome) constitute the components of the indirect effect, i.e., the effect transmitted via the mediator. In practice, the indirect effect will be calculated as a product $\beta_1\gamma_1$.

Of note, the direct effect, i.e., the effect of exposure on the outcome when accounting for the mediator, is different from the total effect, which corresponds the overall effect of exposure on the outcome (denoted as $\alpha_1$ in Figure 4). The total effect captures both the direct and indirect effects and is simply modelled as follows:

$$E[Y|x,c] = \alpha_0 + \alpha_1 x + \alpha_2' c. \quad (3)$$

Similarly to the direct effects, model (3) can be extended for binary responses.

Of note, the familial relatedness between the participants (e.g. G2 siblings) will be taken into account by applying a mixed model based on the genetic correlation matrix.

Causal mediation effects can be estimated from observational data under a set of identification assumptions related to the confounders: (i) no unmeasured exposure-outcome confounders; (ii) no unmeasured exposure-mediator confounders; (iii) no unmeasured mediator-outcome confounders; and

(iv) no mediator-outcome confounders that are causally affected by the exposure. (4) The analysis approach can be extended to cover exposure-mediator interactions and multiple mediators. (5,6)

In practice, to conclude epigenetic inheritance on the basis of mediation analysis, it is first required that the total effect is non-null. In addition, based on the idea of composite null hypothesis for indirect effects, both effects composing the indirect effect need to be present ($\beta_1 \neq 0, \gamma_1 \neq 0$). (7)

In what follows, we first describe how the hypotheses of epigenetic inheritance can be considered as mediation problems. We then describe how the mediators are identified from a large number of epigenetic markers based on epigenome-wide sieving approach (non-coding RNA molecules and DNA methylation markers) and describe how the mediation analysis is applied to estimate the indirect effects via the identified mediators.

*Intergenerational epigenetic inheritance as a mediation problem*

Figure 2 presents two distinct mediation frameworks for intergenerational epigenetic inheritance in two pairs of successive generations, G0–G1 and G1–G2. Intergenerational epigenetic inheritance can be formulated as a mediation question where the effect of paternal (G0 or G1) exposure on the offspring (G1 or G2) phenotype is considered as the total effect, whereas mediation is based on two effects: the effect of paternal exposure on paternal sperm epigenetic markers (i.e., within the G0 or G1 generations) and the effect of paternal sperm epigenetic markers on offspring phenotype. In practice, the total effects are estimated based on model (3) and the mediated effects based on models (1) and (2) after identifying potential mediators among the large number of candidate epigenetic markers (see Sections 4.2).

The observed data will be concluded to support the claim of intergenerational epigenetic inheritance of a specific paternal exposure on a given offspring phenotype if:

1) paternal (G0 or G1) exposure affects the offspring (G1 or G2) phenotype (total effect; Equation (3));

2) epigenetic markers of paternal exposure can be identified (Sections 4.2 and 4.3) ;

3) the identified markers are associated with the offspring phenotype (Sections 4.2 and 4.3);

4) the epigenetic markers identified in both steps 2) and 3) show significant indirect effects in the mediation analysis (Equations (1) and (2)).

A few details in Figure 2 deserve further attention. Firstly, the figure omits showing arrows between phenotypes of different generations. Such between-phenotype associations across generations are assumed to be non-causal and rather based on shared background factors (such as genetics, environment or behaviour, or the preceding causal factors in the DAG). Secondly, there exists a hypothesised arrow from paternal exposure to offspring phenotype via offspring exposure. In the mediation analysis, any potential effects from parental exposure to offspring phenotype via offspring exposure or parental phenotypes are included in the direct effects that incorporate all other potential pathways than the mediation pathway of interest, i.e., the one via epigenetic markers. In addition, while the causal pathways between exposure and sperm epigenetic marker are only emphasised for generations G0 and G1, the effects of exposures on sperm epigenome will be investigated also in the G2 generation when applicable.

*Transgenerational epigenetic inheritance as a mediation problem*
Figure 3 highlights the pathway corresponding to transgenerational epigenetic inheritance over all three generations. Here, the effect of G0 exposure affects the subsequent G1 generation, as in the intergenerational framework, and additionally, the phenotype of G2 generation. Thus, the total effect is the effect of grandpaternal (G0) exposure on the grandchild (G2) phenotype. The mediated effect of exposure is based on the effect of G0 exposure on G0 epigenetic markers, and the effects of G0 epigenetic markers on G2 phenotype. Similarly to intergenerational epigenetic inheritance, the effects are estimated based on models (1)–(3).

The observed data will be concluded to support the claims of transgenerational epigenetic inheritance of a specific grandpaternal exposure on a given grandchild phenotype if:
1) G0 exposure affects the G2 phenotype (total effect; Equation (3));
2) epigenetic markers of G0 exposure can be identified (Sections 4.2 and 4.3) ;
3) the identified markers are associated with the G2 phenotype (Sections 4.2 and 4.3);
4) the epigenetic markers identified in both steps 2) and 3) show significant indirect effects in the mediation analysis (Equations (1) and (2)).

While the G1 generation does not play a role in the direct or indirect effects, the exposure levels of G1 participants need to be taken into consideration; in case a male G1 participant is exposed to the environmental stressor in question, the G2 generation is not only transgenerationally but also inter-generationally exposed. The role of intergenerational exposure via the G1 fathers will be taken into account by primarily examining the G2 offspring of non-exposed G1 parents. It is also important to distinguish the roles of maternal and paternal grandfathers, i.e., examine the transgenerational hypotheses separately for G2 offspring of G1 males and females.

**4.2 Identification of mediators among sperm non-coding RNAs**
To identify potential mediators among the sperm non-coding RNA molecules, two epigenome-wide association studies (EWAS) will be used for each combination of exposure and phenotype: one for the association between (grand)paternal exposure and the (grand)paternal sperm non-coding RNA profile, and one for the association between offspring phenotype and the (grand)paternal sperm non-coding RNA profile. Based on the two EWAS, RNA molecules which after correcting for multiple testing show significant association with both the exposure and offspring phenotype will be considered as potential mediators and used in the mediation analysis.

*Epigenome-wide analyses*
As a pre-processing step for the mediation analysis, an epigenome-wide sieving study will be used to identify non-coding RNA molecules associated with each of the exposures and offspring outcomes. The different non-coding RNA molecule types (piwi-interacting RNA clusters, piwi-interacting RNA sequences, microRNAs and transfer RNAs-derived fragments) will be investigated separately. Of note, at this stage of the analysis, the associations of both exposures and outcomes with the RNA profile will be investigated by treating the ncRNA counts as responses. While for the offspring outcomes these models are not formulated in accordance with the causal order of mediation, they will aid in identifying the RNAs that might play a role between the exposures and responses.

Two different approaches will be used in these EWAS. The effects of exposures on the RNA counts will be investigated using differential abundance analysis based on the negative binomial distribution. (8,9) In addition, to account for the compositional nature of sequencing data sets (10), the read counts within each RNA molecule type will be scaled into relative abundances by dividing the read-count variables with their sum. The effects of exposures on the relative abundances will be assessed using ordinal regression models. For each combination of exposure and response, those RNAs identified to be associated with both exposure and response are carried forward to the analysis of mediation. The epigenome-wide analyses will be run separately for each generation and in addition, when applicable, pooled over the generations.

*Compositional mediation analysis*
After identifying potentially mediating RNAs based on the two EWAS, a hypothesis-driven compositional mediation analysis approach will be used to assess the mediating role of the identified RNAs (11). The relative abundances of the identified RNAs, contrasted to other RNAs, will be transformed into Euclidean coordinates using the *isometric log-ratio transformation* (ilr). RNA molecules belonging to miRNAs, transfer RNA-derived fragments, piRNA clusters, and piRNA sequences will be considered as separate compositions. The ilr-transformations will be built either as *pivotal*, i.e., by contrasting the potentially mediating RNAs against the other RNA molecules within the same group, or based on other *a priori* knowledge of the identified ncRNAs, such as their length or function.

#### 4.3 Identification of mediators in sperm DNA methylation

Similarly to the non-coding RNAs, the potential mediators are identified based on two EWAS as described below. We will use the divide-aggregate composite null test, where the p-values from two EWAS will be combined into one composite p-value based on the distributions of the p-values, utilising the high dimension of the analysis. Based on the composite p-value, potential mediators will be identified after correcting for multiple testing.

*Epigenome-wide analyses*
The effects of exposure on sperm DNA methylation and effect of parental sperm DNA methylation on offspring phenotype will be modelled using EWAS methods for RRBS.

*Mediation analysis*
After candidates for mediating CpG sites have been identified, mediation analysis will be applied to the candidates as described in section *Mediation Analysis*. In case multiple mediators are identified between certain exposures and outcomes, methods allowing for multiple contemporaneous mediators will be utilised. (6,12,13)

#### 4.4 Additional analyses

In addition to the analyses of mediation, the remaining pathways of Figure 1 need to be explored to fully understand the complex interrelationships in multigenerational data. The effects of paternal exposures on offspring sperm epigenetic markers will be investigated by using epigenome-wide association studies similarly as reported above. The similarities in the sperm epigenetic profiles between grandfathers, fathers and offspring will be investigated.

#### 4.5 Additional considerations

*Statistical significance and multiple testing*
Throughout the study, we estimate 95 % confidence intervals and define statistical significance as the interval not containing 0. Within each epigenome-wide analysis, multiple comparisons will be handled using FDR-corrected p-values. Each RNA molecule or CpG site that has an FDR-corrected p-value $< 0.05$ will be considered as a potential mediator and carried forward to the mediation analysis.

*Sensitivity analysis for unmeasured confounding*

Causal inference in mediation analysis is subject to identification assumptions on unmeasured confounding. As the identification assumptions for mediation analysis are untestable, a sensitivity analysis for unmeasured confounders will be used to assess the extent to which the results could have changed if an unmeasured confounder had been taken into account. (14)

*Missing data*

Participants with missing data will be compared to the full analysis set and complete cases. To understand how missing measurements on exposures, mediators or outcomes affects the results, the findings based on complete cases and the full analysis set will be compared.

### REFERENCES


1. Åsenius F, Danson AF, Marzi SJ. DNA methylation in human sperm: a systematic review. Hum Reprod Update. 2020 Nov 1;26(6):841–73.
2. Greeson KW, Crow KMS, Edenfield RC, Easley CA. Inheritance of paternal lifestyles and exposures through sperm DNA methylation. Nat Rev Urol. 2023 Jun;20(6):356–70.
3. Tomar A, Gomez-Velazquez M, Gerlini R, Comas-Armangué G, Makharadze L, Kolbe T, et al. Epigenetic inheritance of diet-induced and sperm-borne mitochondrial RNAs. Nature. 2024 Jun 5;1–8.
4. VanderWeele TJ. Mediation Analysis: A Practitioner's Guide. Annu Rev Public Health. 2016;37(1):17–32.
5. Valeri L, VanderWeele TJ. Mediation analysis allowing for exposure-mediator interactions and causal interpretation: theoretical assumptions and implementation with SAS and SPSS macros. Psychol Methods. 2013 Jun;18(2):137–50.
6. VanderWeele T, Vansteelandt S. Mediation Analysis with Multiple Mediators. Epidemiol Methods [Internet]. 2014 Jan 3 [cited 2024 Jan 12];2(1). Available from: https://www.degruyter.com/document/doi/10.1515/em-2012-0010/html
7. Liu Z, Shen J, Barfield R, Schwartz J, Baccarelli AA, Lin X. MEDIATION x Large-Scale Hypothesis Testing for Causal Mediation Effects with Applications in Genome-wide Epigenetic Studies. J Am Stat Assoc. 2022 Jan 2;117(537):67–81.
8. Love MI, Huber W, Anders S. Moderated estimation of fold change and dispersion for RNA-seq data with DESeq2. Genome Biol. 2014 Dec 5;15(12):550.
9. Robinson MD, McCarthy DJ, Smyth GK. edgeR: a Bioconductor package for differential expression analysis of digital gene expression data. Bioinformatics. 2010 Jan 1;26(1):139–40.
10. Gloor GB, Macklaim JM, Pawlowsky-Glahn V, Egozcue JJ. Microbiome Datasets Are Compositional: And This Is Not Optional. Front Microbiol. 2017;8:2224.
11. Kartiosuo N, Nevalainen J, Raitakari O, Pahkala K, Auranen K. Hypothesis-driven mediation analysis for compositional data: an application to gut microbiome. Biostat Epidemiol. 2024 Jan 2;8(1):e2360375.
12. Kim C, Daniels MJ, Hogan JW, Choirat C, Zigler CM. MEDIATION BAYESIAN METHODS FOR MULTIPLE MEDIATORS: RELATING PRINCIPAL STRATIFICATION AND CAUSAL MEDIATION IN THE ANALYSIS OF POWER PLANT EMISSION CONTROLS. Ann Appl Stat. 2019 Sep;13(3):1927–56.
13. Wang W, Nelson S, Albert JM. Estimation of causal mediation effects for a dichotomous outcome in multiple-mediator models using the mediation formula. Stat Med. 2013 Oct 30;32(24):4211–28.
14. Imai K, Yamamoto T. Identification and Sensitivity Analysis for Multiple Causal Mechanisms: Revisiting Evidence from Framing Experiments. Polit Anal. 2013;21(2):141–71.
